# COVID-19 Transmission Dynamics Underlying Epidemic Waves in Kenya

**DOI:** 10.1101/2021.06.17.21259100

**Authors:** Samuel P. C. Brand, John Ojal, Rabia Aziza, Vincent Were, Emelda A Okiro, Ivy K Kombe, Caroline Mburu, Morris Ogero, Ambrose Agweyu, George M Warimwe, James Nyagwange, Henry Karanja, John N Gitonga, Daisy Mugo, Sophie Uyoga, Ifedayo M O Adetifa, J Anthony G Scott, Edward Otieno, Nickson Murunga, Mark Otiende, Lynette I Ochola-Oyier, Charles N Agoti, George Githinji, Kadondi Kasera, Patrick Amoth, Mercy Mwangangi, Rashid Aman, Wangari Ng’ang’a, Benjamin Tsofa, Philip Bejon, Matt. J. Keeling, D. James. Nokes, Edwine Barasa

**Affiliations:** Kenya Medical Research Institute (KEMRI) -Wellcome Trust Research Programme (KWTRP), Kilifi, Kenya; The Zeeman Institute for Systems Biology and Infectious Disease Epidemiology Research (SBIDER), University of Warwick, UK; School of Life Sciences, University of Warwick, UK; London School of Hygiene and Tropical Medicine (LSHTM), UK; Health Economics Research Unit, KEMRI-Wellcome Trust Research Programme, Nairobi, Kenya; Population Health Unit, Kenya Medical Research Institute -Wellcome Trust Research Programme, Nairobi, Kenya; Centre for Tropical Medicine and Global Health, Nuffield Department of Medicine, University of Oxford, Oxford, United Kingdom; Department of Infectious Diseases Epidemiology, London School of Hygiene and Tropical Medicine, London, United Kingdom; Ministry of Health, Government of Kenya, Nairobi, Kenya; Presidential Policy & Strategy Unit, The Presidency, Government of Kenya; Mathematics Institute, University of Warwick, UK

**Author notes:** Correspondence to: Samuel Brand and/or John Ojal. Samuel Brand and John Ojal contributed equally.

## Abstract

Policy decisions on COVID-19 interventions should be informed by a local, regional and national understanding of SARS-CoV-2 transmission. Epidemic waves may result when restrictions are lifted or poorly adhered to, variants with new phenotypic properties successfully invade, or when infection spreads to susceptible sub-populations. Three COVID-19 epidemic waves have been observed in Kenya. Using a mechanistic mathematical model we explain the first two distinct waves by differences in contact rates in high and low social-economic groups, and the third wave by the introduction of a new higher-transmissibility variant. Reopening schools led to a minor increase in transmission between the second and third waves. Our predictions of current population exposure in Kenya (∼75% June 1st) have implications for a fourth wave and future control strategies.

**One Sentence Summary:** COVID-19 spread in Kenya is explained by mixing heterogeneity and a variant less constrained by high population exposure

## Main text

Following the first PCR confirmed case of COVID-19 in Kenya on 13^th^ March 2020, the Kenyan government rapidly introduced measures aimed at suppressing SARS-CoV-2 transmission in the country. These measures included: the closure of international borders, with the exception of cargo movement; closing of schools and other learning institutions; a ban on social gatherings and meetings; closure of places of worship, bars and restaurants; a dawn to dusk curfew; mandatory wearing of masks in public places; physical distancing guidelines including on public transportation; and restrictions on movement into or out of counties with high infection rates including the two main Kenyan cities, Nairobi and Mombasa (*1*)(Fig. 1). Despite these measures the rate of new COVID-19 cases grew in Kenya indicating that measures had not been enough to consistently push the effective reproduction number R(t) < 1. Moreover, serological surveillance indicated that a higher than expected fraction of the Kenyan population had been exposed to SARS-CoV-2 given the case reports at the time: June 2020 adjusted seroprevalences, based on blood donor samples from the Kenya National Blood Transfusion Services (KNBTS), were 5.6% for Kenya, 8% for Mombasa, and 7.3% for Nairobi (*2*).

**Figure 1:**
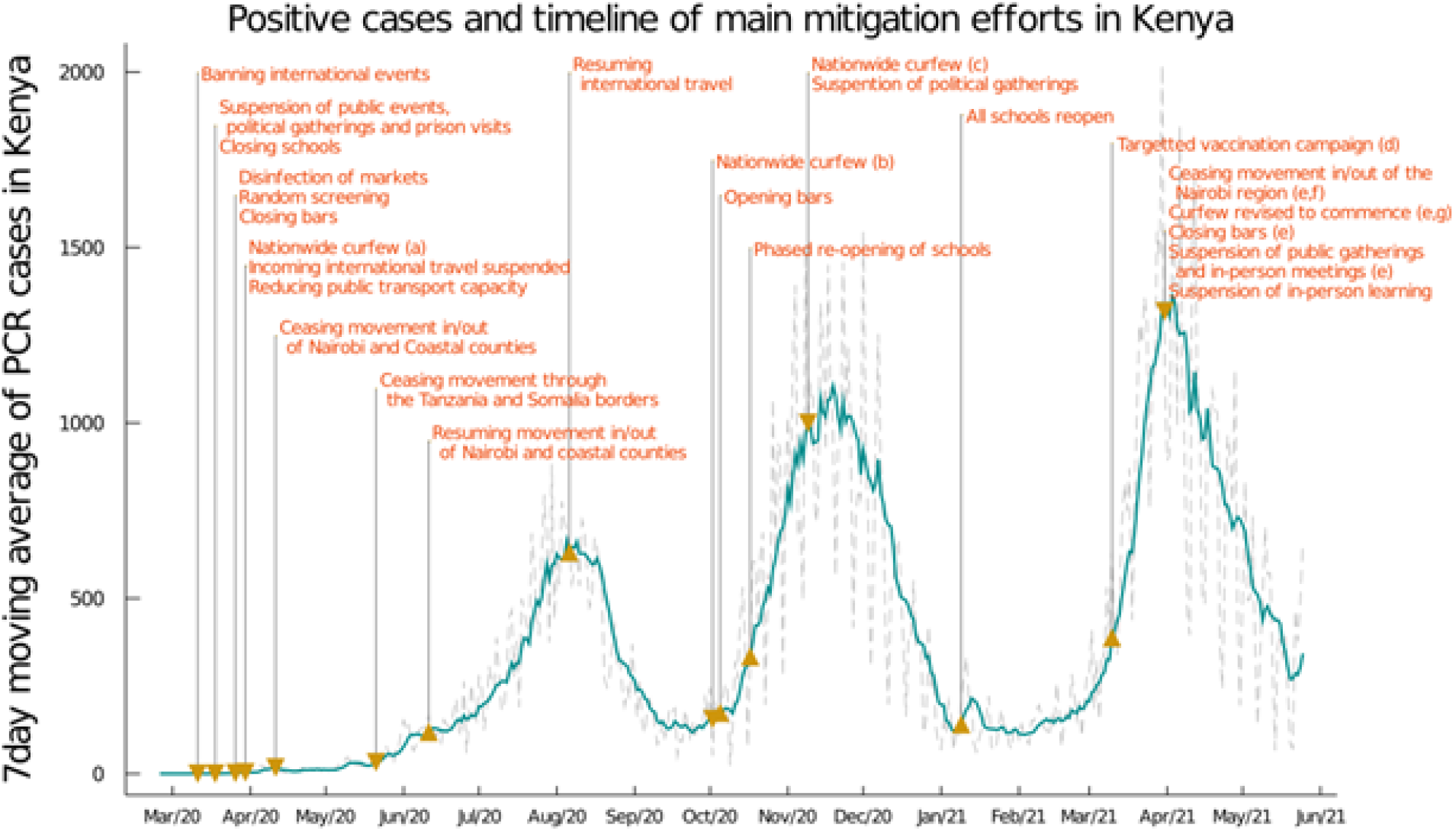
7day moving average of daily positive PCR tests from the Kenyan national linelist and a timeline of the main mitigation events applied by the Kenyan government representing tightening (down-arrow) and relaxation (up-arrow) of measures, with: (a) curfew from 7pm to 5am; (b) curfew from 11pm to 4am; (c) curfew from 10pm to 4am; (d) front line workers and individuals older than 58 years (approximately 1.2m doses); (e) the region includes Nairobi, Kajiado, Machakos, Kiambu, Nakuru; (f) this restricted movement into and out of the block of counties in (e) but not between these counties; (g) curfew from 8pm to 4am.

Detected COVID-19 incidence in Kenya first peaked in early August 2020 during a period of relaxation of measures: the end of the Nairobi and Coastal counties (including Mombasa) lockdown (7^th^ June 2020), and the resumption of international air travel (1^st^ August 2020). A single-wave epidemic in Kenya peaking within 100-200 days after SARS-CoV-2 introduction into the country was initially predicted, based on assumptions that included a single population group, and the development of immunity to reinfection (*3-6*). However, second and third waves occurred in mid-November 2020 and in March 2021, respectively. Multiple waves of COVID-19 incidence in High Income Country (HIC) settings have usually been associated either with a relaxation of previous restrictions, or establishment of the epidemic in a new geographical area. More recently, the emergence of new variants has been associated with further waves of infection (*7*). In Kenya, and other countries in Africa, a temporal association between relaxation of restrictions and subsequent waves is implausible. Understanding the causation of such multiple waves is critical for forecasting hospitalization demand and the likely effectiveness of interventions including vaccination strategy.

There are multiple potential explanations for sequential wave dynamics in COVID-19 incidence, which might be acting singly or in concert: social clustering (*8*), changing adherence to measures over time (*9*), seasonal effects on transmission (*10*), re-opening of places of learning (*11*), lower transmission rates in rural settings leading to later peaks in those areas (*4*), waning immunity after an infection episode, as well as the introduction of new SARS-CoV-2 variants which are more transmissible than previous strains, or/and, evade prior immunity acquired by natural infection (*12*). In this work, we present evidence that the most plausible explanation for the pattern of cases and seroprevalence observed in Kenya is a combination of differential adherence to measures between sub-populations which we identify with lower and higher socio-economic status (SES) in 2020 followed by a sharp increase in virus transmissibility in 2021, consistent with that observed in other countries affected by variants of concern, e.g. the United Kingdom (*13*) and India (*14*).

We developed a county-specific, two-socio-economic status (SES) group, SEIRS-type transmission model, using a waning immunity rate derived from recent studies on the protectiveness of a natural infection to future reinfection (*15-18*), which was parametrised using PCR test data derived from the Kenyan national COVID-19 database, Google data on smartphone mobility and serological test data from the on-going longitudinal serosurveillance using samples from KNBTS donors (*2*). We used a hierarchical approach to inferring the underlying epidemic trajectories in each of the 47 Kenyan semi-autonomous counties by the following three steps: a) grouping counties by similarity over a range of sociological and epidemiological metrics using machine learning; b) for the 11 counties with a relatively high density of serology tests we jointly inferred epidemiological model parameters e.g. i) baseline R0 for the county, ii) the effect of schools being open on R(t), iii) the increase in transmissibility in February 2021 when B.1.1.7 lineage (alpha variant) SARS-CoV-2 was first detected in Kenya (*19*), iv) the fraction of the population in the higher SES group in each county and their assortative mixing rate, and v) the fraction of cases reported for the county using Hamiltonian Markov chain Monte Carlo (*20*) with mildly informative priors, and c) we inferred model parameters for the remaining 36 counties using informative priors for reporting fractions derived from a synthesis of the posterior distributions of counties grouped as similar to that county (see *Supporting information* for details).

The two-SES group transmission model was able to capture the timing and intensity of all three waves of Kenyan COVID case incidence and the trend of increasing proportion seropositive among KNBTS donors (Fig. 2). The model fits estimate that the high SES group, which we characterized by higher likelihood of smartphone ownership, had a reduction in movement after initial restrictions were announced with delayed reversion to pre-pandemic movement patterns and a higher likelihood of being tested and being admitted to hospital. The lower SES group, who were unlikely to own smartphones, did not reduce movement with initial restrictions to the same degree and reverted to baseline rates sooner, relative to the higher SES group, and were less likely to be tested or admitted to hospital (see *Supporting information* for predicted mixing rates for the upper SES group). We also validated the fitted model by comparing forecasts of seropositivity rates with those from data not used to infer model parameters. We used rounds 1 and 2 of the seropositivity survey using KNBTS donors for model parameter inference, collected during May – September 2020. Estimated seroprevalence among the Kenyan population, derived from the fitted two-SES group transmission model, was in good agreement with the out-of-sample round 3 of KNBTS seroprevalence data, collected January – March 2021 (Fig. 2). The Nairobi-specific epidemic trajectory inferred in this study agrees with seroprevalence estimates from a randomised survey from Nairobi, and, is congruent with the observation that it was public hospitals in Nairobi (favoured by lower SES groups) that came under pressure in the first wave, whereas the second wave showed increased admission to private health facilities (Fig. S7-8). As well as capturing the past trends of case reporting and seropositivity in Kenya, the fitted two-SES group transmission model accurately predicts the daily rate of new confirmed COVID-19 cases reported by the Kenyan Ministry of Health for the month after the censoring date of the PCR test data used to infer model parameters (Fig. 2). This contrasts the one-group model which provided no plausible explanation of the second wave (see *supporting information*).

**Figure 2:**
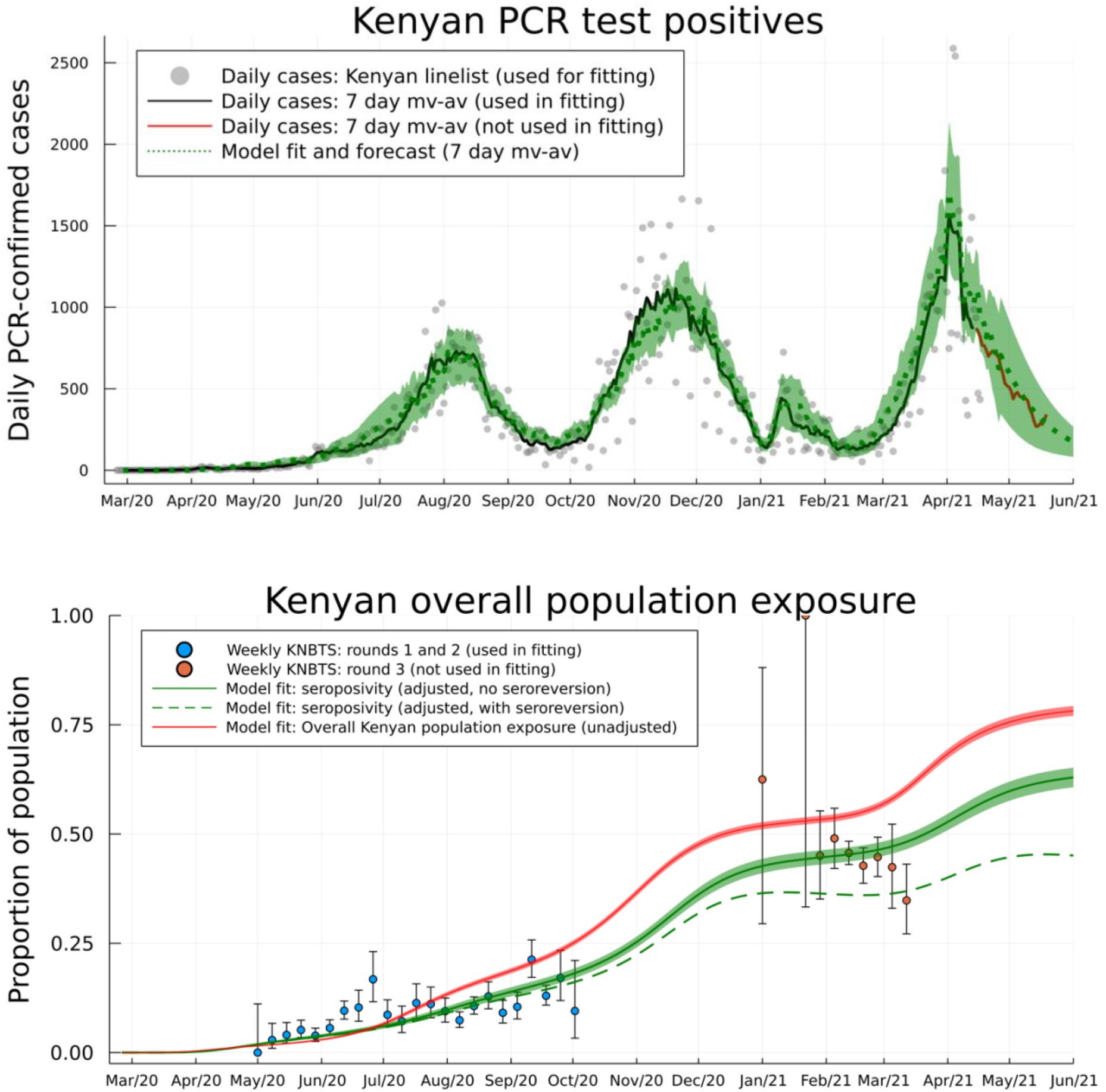
Daily PCR confirmed COVID-19 cases (*top*) and weekly serology estimates from KNBTS donors with overall attack rate estimates (*bottom*). Shown are daily numbers of PCR test positives from the Kenyan national linelist (*top*; grey dots are daily reports used for fitting the model, curves are 7-day moving averages). The model prediction for the 7-day moving average of daily case incidence (*top;* green dash curve, shading shows 3-sigma intervals) were derived from inference on the county-specific linelist PCR data and rounds 1 and 2 of the KNBTS serology survey (*bottom;* blue dots). Predictions before mid-April 2021 are back-calculations using known numbers of PCR tests per day, whereas, after mid-April 2021 model predictions are forecasts which also estimate the number of PCR tests that will occur per day in each county. We show the next month of PCR test positive data, not used in fitting, as a validation of the model short-term predictive accuracy (*top;* red curve). Back-calculated model estimates of seropositivity (*bottom;* green solid curve) are shown with round 3 of the KNBTS serology survey data (*bottom;* red dots, not used in model inference). We also show back-calculated estimates of seropositivity under the assumption that median time to seroreversion (loss of detectable antibodies rather than loss of immunity) from infection was one year. Model estimates of overall Kenyan seropositivity are adjusted from county-specific estimates by weighting by number of serology tests in each county (over KNBTS rounds 1 and 2). The overall estimated Kenyan attack rate (population exposure) is shown as unadjusted (*bottom*; red curve).

The two-SES group transmission model reconciled the apparent paradox between evidence of the effectiveness of the rapidly introduced Kenyan measures in reducing mobility out of the home among Kenyan smartphone users, which was close to that observed in European and North American countries (Fig. S1), and that case rates and fatality rates display two distinct waves in Kenya in 2020. In some Kenyan counties (e.g. the urban counties Nairobi and Mombasa, and some of the semi-urban counties) we infer that a substantial group of people belong to the higher SES group whose mobility is well-represented by Google smartphone data; a combination of school closures and reduction in mobility (by 44.5% see *supporting information*) reduced the effective reproductive number sufficiently that newly infected people among the higher SES group were on average generating less than 1 secondary infection by April 2020 (Fig. 3). The growth rate in cases, and relatively high levels of seroprevalence among KNBTS donors, are explained by the rest of the population in the lower SES group having R(t) > 1 into May and June 2020 (Fig. 3). The model inferred that the reduction in mobility among the lower SES group was substantially less than among the higher SES group: the posterior mean estimate for reduction in mobility among the lower SES group in Nairobi was 13.8% (CI 11.3-17.5%), in Kenya’s second city Mombasa was 18.9% (CI 17.4-20.4%), and posterior mean estimates for lower SES group mobility reduction across all 47 Kenyan counties had a median of 15.7% (IQR 10.9-19.6%). We assumed that school closures reduced R(t) for both SES groups equally. The inferred reduction in R(t) due to schools closing varied from county to county, the median reduction in R(t) over counties was 13.5% (IQR 4.3-23.7%; Nairobi estimate for school closure effect was 23.8% CI 16.5-31.6%, Mombasa estimate for school closure effect was 20.2% CI 15.2-25.2%; Fig. 3).

**Figure 3:**
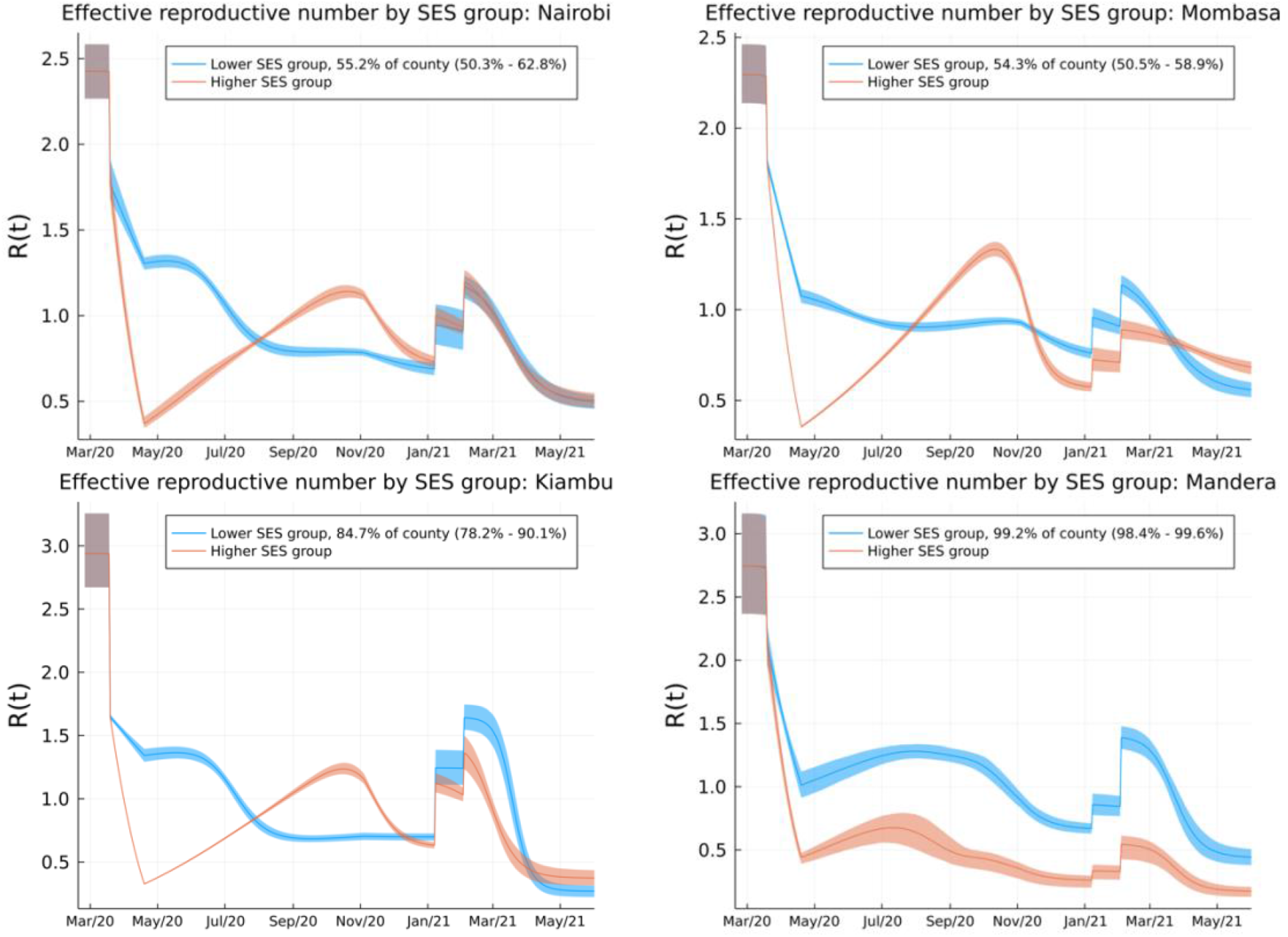
Effective reproductive number over time (R(t)) for lower and higher SES groups in four representative counties: Nairobi (*top left*), Mombasa (*top right*), Kiambu (*bottom left*), and, Mandera (*bottom right*). Nairobi and Mombasa are Kenya’s two largest cities and form fully urban counties, Mandera county has a largely rural population and is remote from the main urban conurbations, Kiambu county borders Nairobi and has a ∼60% urban population. The transmission model infers the proportion of the population in each SES group in each county. The highest proportion of higher SES group individuals are inferred to be in Nairobi and Mombasa out of all counties, whereas for Mandera county very close to all individuals are inferred as being in the lower SES group and the model effectively defaults to one group SEIRS transmission. The model inference for R(t) in Kiambu represents a county between these two extremes. In each county, the first discontinuous increase in R(t) is due to schools reopening, and the second is due to more transmissible variants becoming dominant in transmission.

The second wave in Kenya in 2020 was triggered by the higher SES group returning to pre-COVID-19 mobility patterns by early November 2020 (Fig. S1), and, therefore, R(t) going above 1 for the higher SES group (Fig. 3). Low rates of mobility somewhat shielded the higher SES group from infection in the first wave among the lower SES group. Therefore, the lower SES group suffered two waves in 2020, whereas the higher SES group effectively suffered one wave peaking in late 2020 (Fig. 4). The overall detection rate was determined in part by the number of PCR tests performed each day, and this rate dropped in September 2020 (Fig. S4). A consequence of the drop in the testing rate was that the case reporting shows a much sharper delineation between the first two waves (Fig. 2) than the underlying inferred infection rates (Fig. 4), which reveal that there was only a moderate dip in infections in August/September 2020. By accounting for the delay between infection and COVID-19 fatality, and fitting SES group specific infection-fatality-detection ratios (IFR-detection, see *methods and supporting information*) to each county, we found reasonable agreement between the predicted and observed timings of peak fatality rates in Kenya (Fig. 4). Overall, our model-based estimate was that only 11% of the total Kenyan population were in the higher SES group, whose mobility was well-described by Google mobility data, with the highest concentration of higher SES group individuals in the two main cities: 43.4% of the Nairobi population (CI 35.4-49.2%) and 40.3% of the Mombasa population (CI 35.0-45.4%). Additionally, we estimate that infections among the higher SES group were substantially more likely to be detected than among the lower SES group: odds ratio for Nairobi 4.5 (CI 1.5-17.9), for Mombasa 4.8 (CI 3.2-6.8). The odds ratio between detection per infection in the two SES groups was inferred to be even more extreme across Kenya as a whole, with substantial variation from county to county: median odds ratio estimate over counties was 18.5 (counties estimate IQR 2.5-27.9), although most counties had a small number of people in the higher SES group.

**Figure 4:**
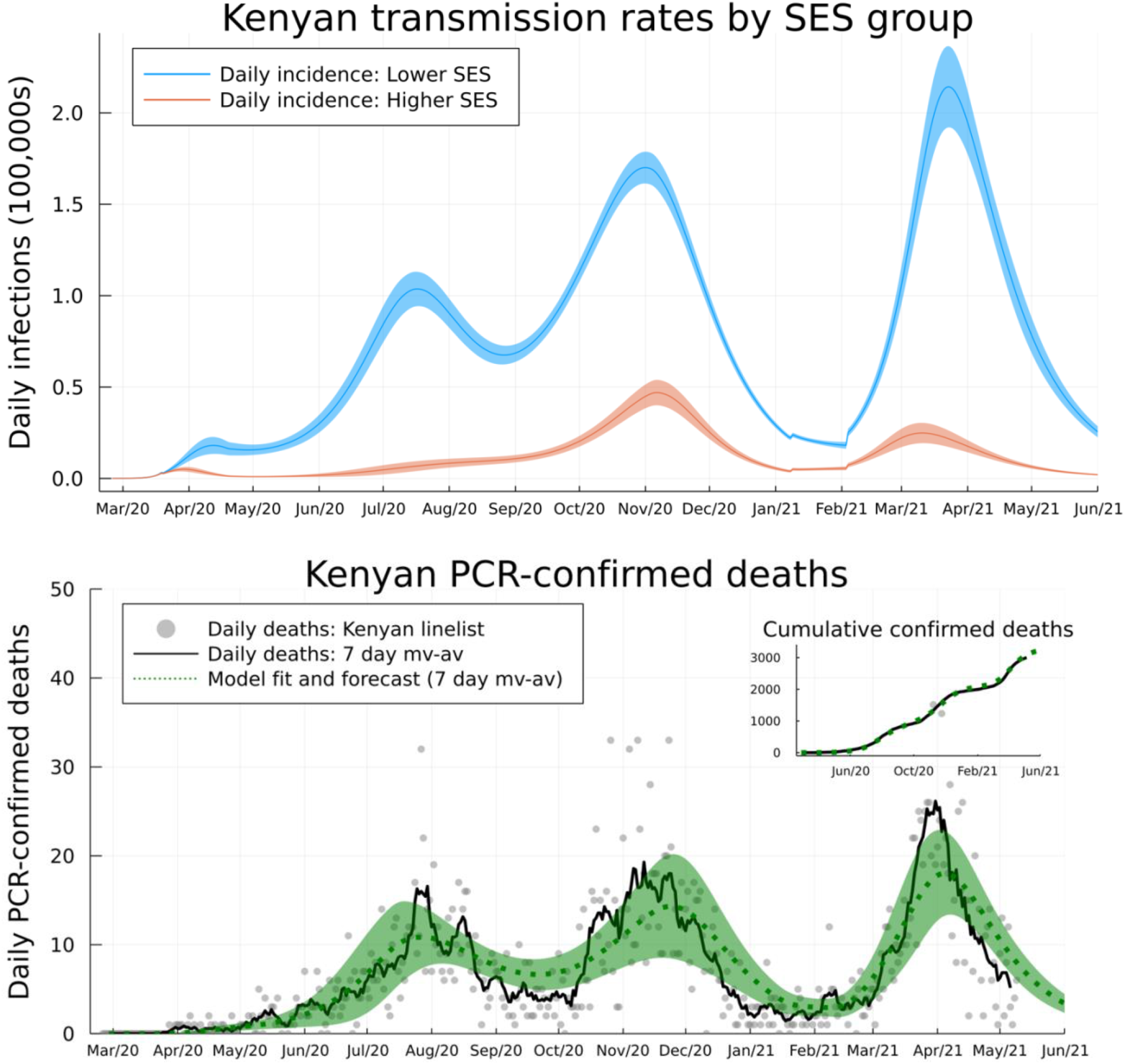
Model inferred underlying true incidence rates by SES group relative to the whole Kenyan population size (*top*) and reported PCR-confirmed deaths due to COVID-19 disease (*bottom*). The size of the upper SES group was estimated to be 11% of the Kenyan population, explaining the lower absolute rate of incidence (red curve) compared to the lower SES group (blue curve). We inferred that the lower SES group have experienced three waves of SARS-CoV-2 transmission, whereas the upper SES group has experienced two. The model fit for seven day moving average (green dashed curve, with shading as 95% PIs) is shown against the seven-day moving average for deaths reported in the Kenyan linelist (black curve). Cumulative observed and fitted deaths are shown in the top-right inset.

Fully reopening schools in January 2021 was associated with a slight increase in cases and deaths in Kenya, with a peak in January and early February 2021 (Figs. 3,4). However, school reopening does not explain the third wave in Kenya observed in March and April 2021, which was considerably larger than the increase in January/February 2021. The two SES group model was not a sufficient explanation for a third wave, neither was loss of immunity or any detectable trend in mobility. The first cases of the more transmissible Alpha variant B.1.1.7 were identified in Kenya from mid-January 2021(*19*). We investigated if the data supported an increase in transmissibility per infected person starting at the beginning of February 2021 as well as an influx of new exposed individuals representing domination of wildtype strains of SARS-CoV-2 by a fitter new variant. In the Kenyan urban counties, we found credible range of increase in transmissibility of 15.0-46.6% (Nairobi 32.5% CI 18.1-46.6%; Mombasa 22.8% CI 15.0-31.2%), and this was reflected in increased transmissibility estimates across Kenyan counties: median over county estimates 46.1% (IQR 31.6-72.9%). The fitted model predicted that this large increase in transmissibility will push the overall exposure to SARS-CoV-2 in Kenya from a back-calculated estimate of 53.5% in February 2021 to 78.1% by June 2021 (Fig. 2). The rate of seroreversion, that is the loss of detectable antibodies rather than necessarily loss of protective immunity, has been identified as an important quantity for estimating population exposure prevalence from serological data (*21*). Because the serological data used for parameter inference was collected within 7 months of the first identified case in Kenya, we assumed that seroreversion was negligible over this period. However, we note that assuming no future seroreversion led to closer agreement between model back-calculation and round 3 KNBTS data than assuming a median 1 year between infection and seroreversion (Fig. 2); that is that our modelling doesn’t support the need to incorporate seroreversion to capture the true population exposure over the time scale of a year, unlike for Buss *et al* (*21*). This finding highlights that seroreversion rate depends on the serological assay used (*22*) and cannot necessarily be extrapolated between settings. A negligible seroreversion rate may be more applicable for the ELISA used in Kenya where the cut-offs prioritize specificity over sensitivity (*2, 23*).

Our modelling exercise provides a credible mechanistic interpretation of the three waves of COVID-19 in Kenya. To do this, we invoke two key underlying assumptions. First, a stratified population differing in mobility (associated with lower and higher SES), and second, increased virus transmissibility compatible with competitive succession of a SARS-CoV-2 variant of concern in wave 3. A key simplifying assumption in this modelling study is that we assumed that the diversity of behaviours across the population in each Kenyan county can be reduced to stratifying into two groups with assortative mixing favouring transmission within group, and identifying these groups into lower and higher SES groups based on similarity to mobility trends among smart phone users. We believe that this is a well-evidenced hypothesis for Kenya.

Marked social and economic structuring has been described in Kenya; 36% of the population live below the national poverty line (*24*) and 55% live in informal settlements (*25*). Further, 83% of Kenya’s labour market is informal, characterized by unstable and unpredictable daily wages (*26*). Lower socio-economic groups have been identified as vulnerable to SARS-CoV-2 in the global South due to residence in informal settlements at high population density, reduced access to sanitation, and dependence on informal employment requiring daily mobility (*27, 28*). In contrast, the higher SES group with job security can work from home, socially distance and readily access water and sanitation, thereby decreasing transmission. In Kenya, Google mobility data from smartphone users indicates a sharp decline in movement to settings outside of the home (Fig. S1). We found that the two SES group model used in this paper was able to capture pattern of cases and seroprevalence in Kenya over the first three waves, despite radically simplifying the underlying social structure.

We predict the proportion of the Kenya population exposed to SARS-CoV-2 to be greater than 75% by the beginning of June 2021 (Fig.2), corresponding to around 39 million people. However, less than 4,000 confirmed COVID-19 deaths and 180,000 confirmed SARS-CoV-2 infections have been identified as of the 1^st^ June 2021. We found that people among the lower SES group were likely to be even more under-sampled than people among the upper SES group, as well as identifying wide regional variation in the detection rate. These results emphasize the necessity of community surveys of SARS-CoV-2 prevalence, exposure, and an investigation into the under-reporting of mortality and severe disease due to COVID-19.

The high population exposure suggests that a fourth COVID-19 wave in Kenya before the end of 2021 would only be likely if (i) a variant arises with substantial further enhancement in transmissibility or immune escape, such as the B.1.617.2 Delta variant (*29*), or (ii) significant waning of immunity in those previously exposed. Nevertheless, the predicted geographical (i.e. county) variation in the proportion exposed will likely result in an extended tail to the current epidemic wave. We conclude that our analysis which triangulates PCR test, seroprevalence, mobility and genomic data to develop a coherent explanation of the transmission dynamics of COVID-19, provides insight of public health importance in Kenya, including targeting health care capacity and pharmaceutical and non-pharmaceutical interventions.

## Data Availability

All code and data for the transmission model underlying this study is accessible at the github repository https://github.com/SamuelBrand1/kenya-covid-three-waves. All data is available in the main text or the supplementary materials.

## Acknowledgments

We thank staff of the Kenya National Blood Transfusion Service and the blood donors, the county Rapid Response Teams and SARS-CoV-2 PCR testing laboratories, and data management staff of the Ministry of Health, Government of Kenya. This paper is published with permission of the Director of KEMRI.

## Author contributions

**Brand SPC**: Conceptualization, Formal Analysis, Methodology, Software, Writing – Original Draft Preparation, Writing – Review & Editing;

**Ojal J**: Conceptualization, Formal Analysis, Methodology, Software, Writing – Original Draft Preparation, Writing – Review & Editing;

**Aziza R**: Conceptualisation, Formal Analysis, Methodology, Project Administration, Software, Writing – Review & Editing;

**Were V**: Conceptualization, Writing – Review & Editing;

**Okiro EA**: Data Curation, Supervision, Writing – Review & Editing;

**Kombe IK**: Formal Analysis, Methodology, Software, Writing – Review & Editing;

**Mburu C**: Formal Analysis, Methodology, Software, Writing – Review & Editing;

**Ogero M**: Data Curation, Writing – Review & Editing;

**Agweyu A**: Conceptualization, Project Administration, Funding Acquisition, Writing – Review & Editing;

**Warimwe GM**: Conceptualization, Supervision, Funding Acquisition, Writing – Writing – Review & Editing

**Nyagwange J, Karanja H, Gitonga JN, Mugo D:** Serology methodology,

**Uyoga S**: Data Curation, Investigation, Writing – Review & Editing;

**Adetifa IMO**: Conceptualization, Supervision, Writing – Review & Editing;

**Scott JAG**: Conceptualization, Supervision, Funding Acquisition, Writing – Review & Editing;

**Otieno E, Otiende M, Murunga N**: Data Curation, Writing – Review & Editing;

**Ochola-Oyier LI**: Data Curation, Investigation, Writing – Review & Editing;

**Agoti CN, Githingi G**: Data Curation, Investigation, Writing – Review & Editing;

**Kasera K**: Project Administration, Writing – Review & Editing;

**Amoth P**: Project Administration, Writing – Review & Editing;

**Mwangangi M**: Project Administration, Writing – Review & Editing;

**Aman R**: Project Administration, Writing – Review & Editing;

**Ng’ang’a W**: Project Administration, Writing – Review & Editing;

**Tsofa B**: Project Administration, Writing – Review & Editing;

**Bejon P**: Conceptualization, Writing – Original Draft Preparation, Writing – Review & Editing;

**Keeling MJ**: Conceptualization, Formal Analysis, Funding Acquisition, Methodology, Software, Supervision, Writing – Original Draft Preparation, Writing – Review & Editing;

**Nokes DJ**: Conceptualization, Funding Acquisition, Project Administration, Supervision, Writing – Original Draft Preparation, Writing – Review & Editing

**Barasa E**: Conceptualization, Project Administration, Writing – Original Draft Preparation, Writing – Review & Editing;

