## supporting_information_text for "COVID-19 Transmission Dynamics Underlying Epidemic Waves in Kenya"

**This document includes:**

Materials and Methods
Supplementary Text
Figs. S1 to S8
Tables S1
Captions for Data S1 to S7

**Other Supplementary Materials for this manuscript include the following:**

Data S1 to S4

### Materials and Methods

#### Data description

In this paper, we make inferences of the penetration of SARS-CoV-2 transmission into each Kenyan county using a mechanistic transmission model. Joint posterior distributions for the parameters of the transmission model were inferred for each county using a synthesis of three data sources:

- **Kenyan Ministry of Health National linelist.** We were provided linelist information about confirmed cases and swab tests performed. However, the available data differed by date and county. Below we describe which bits of data were available at different times:
    - Confirmed cases. PCR positive swab samples by collection date, $(ObsP_1^+)_n$, denoting the number of positive test results collected on day *n* for each county,, using the lab confirmation date of the swab test. Negative tests were not recorded in the national linelist. Those confirmed cases who died were also recorded by date of death for each county, $(X^+)_n$. The national linelist case data was available over the period we used for parameter inference in this paper (13th March to 26th April 2021), and over the period we used for out-of-sample validation of the model's predictive ability (27th April – 21st May 2021). We screened out tests due to contact tracing or at entry to Kenya.
    - National combined laboratory test results. The combined reported tests, by laboratory collection date, across Kenyan laboratories, $(ObsP_2^+)_n$ and $(ObsP_2^-)_n$, respectively denote the positive and negative test results by confirmation date among all Kenyan laboratories. This data covered the period 16th June onwards for all counties.
    - Kemri-Wellcome Trust Research Programme (KWTRP) test result linelist. Most swab tests performed in the counties of the Coastal Province of Kenya reverted to the KWTRP testing laboratory for confirmation. $(ObsP_3^+)_{n,c}$ and $(ObsP_3^-)_{n,c}$, respectively denote the positive and negative test results by collection date in coastal county *c* on day *n* that were confirmed at KWTRP. As with the national linelist we screened out tests due to contact tracing or at entry to Kenya. The KWTRP linelist data was available from 13th March onwards. The counties covered by the KWTRP linelist were Kilifi, Kwale, Lamu, Mombasa, Taita Taveta and Tana River.

- **KWTRP serological surveillance programme (rounds 1 and 2).** Numbers of seropositive $(ObsS^+)_{n,c}$and seronegative $(ObsS^-)_{n,c}$ blood samples collected from regional centres of the Kenyan National Blood Transfusion Service (KNBTS) on day *n* originating from county *c*. Residual blood samples for serology were obtained from regular blood donors attending 4 regional KNBTS centres (Mombasa, Nairobi, Eldoret and Kisumu) in two rounds of collection in May – September 2020. The study methodology is fully described in Uyoga et al (*2*). We also show data from round 3 of the KWTRP serological surveillance programme (unpublished) in the main manuscript Fig.

2, which was collected January – March 2021, as validation data for the model predictive ability of seroprevalence.

- **Google mobility data.** Daily estimates of relative human mobility $m_{n,area}$ compared to a baseline of the same date in the previous year (2019) derived from Google mobility trends (*30*). We assumed that changes in trends in SARS-CoV-2 transmission in Kenya were due to changes in the underlying population mobility. In particular, by changing frequency of indoor congregations. Therefore, we calculated $m_{n,area}$ as the average change in baseline mobility over the “retail and recreation”, “grocery and pharmacy”, “transit stations”, and, “workplaces” settings (Google defined categories), and also over the week prior to day *n*, in order to average over weekend effects. Due to incomplete data, and the likely bias introduced by using a mobility estimate derived from smartphone users in predicting the mobility of semi-rural populations outside of the major urban conurbations in Kenya, we consider only three areas: Nairobi, Mombasa and the pan-Kenyan aggregate (Fig. S1).

We performed a data cleansing process on the PCR datasets to improve their quality regarding the test date, age of the individual, symptoms and location. The data contained 4 spatial attributes; county, sub-county, ward and village, which we correct for inaccuracies in line with a ward level spatial baseline which accounts for the 47 Kenyan counties, 295 sub-counties and 1450 wards (https://data.humdata.org/dataset/administrative-wards-in-kenya-1450). This process includes spelling checks, string distance calculations, and an automated geo-search of addresses constructed using different combinations of the available spatial data per record. Finally, we run a distributed script that allows collating the number of positive/negative tests per day, county, sub-county, and age.

For making inference about unknown parameters for each county (see below) we used different sources of PCR swab testing data depending on the county and date according to the following rules:

- For coastal counties the KWTRP linelist was used.
- For non-coastal counties the national case linelist was used between 13th March and 6th July, negative test results are unavailable over this period.
- For non-coastal counties the national laboratory test linelist was used after 7th July and 21st May 2021. During this period both positive and negative test results were available.

The combined linelist data used in this paper is provided (Data S2), as well as the daily serology samples (Data S3).

#### Transmission model

We model the spread of SARS-CoV-2 in each of the 47 Kenyan counties as a two group SEIRS transmission process, with waning immunity returning completely protected recovered individuals to a waned immunity partially protected state (W). We attempt to capture the basic features of transmission between households, that is that we focus on the reduction in mixing indoors settings outside the home (termed places-of-interest/POIs) due to changes in behaviour in response to Kenyan government measures and an increased sense of personal risk. This approach is conceptually similar to some modelling studies in high-income countries (e.g. (*31*)), however, we used a simpler model structure reflecting the lower resolution data available in Kenya compared to most HICs.

We hypothesize that social disaggregation occurred in Kenyan counties; that is that there were broadly two groups of people in each county who differed in their ability to reduce social mobility. These two groups represented the people with lower and higher socio-economic status (SES) in each county. Both the higher and lower SES groups are assumed to have similar epidemiological characteristics except that we assumed that:

- The lower and higher SES groups responded to Kenyan government measures by reducing their time spent in POIs at different rates.
- The higher SES group reduction in time spent in POIs was well estimated by Google mobility trend data (*30*).

Alternatively, we can think of the two groups as being *defined* as those people whose visitation rates to POIs are well represented by Google mobility data (an unknown sized group in each county), and since this requires ownership of a smartphone, estimated as 30% of Kenyan adults in 2017 (*32*), this definition selects for being in a higher socio-economic status.

We assumed that transmission is sustained by mixing at POIs and, therefore, if the SES groups reduced their mixing in POIs by proportions, respectively, $c_L(t)$ and $c_U(t)$, at each time *t* then the reproductive number for each county was reduced from some baseline reproductive ratio $R_0$ that would have be realised if social mobility had remained unchanged. By assumption, $c_U(t)$ was fitted to Google mobility data, whereas $c_L(t)$ was inferred jointly with other model parameters (see below). The per capita force of infection on individuals in the lower and higher SES groups, denoted respectively $\lambda_L(t)$ and $\lambda_U(t)$, was

$$\begin{aligned} \lambda_L(t) &= \frac{\gamma R_0(t)}{N_L}\big(\epsilon c_L(t) I_L(t) + (1-\epsilon) c_U(t) I_U(t)\big), \\ \lambda_U(t) &= \frac{\gamma R_0(t)}{N_U}\big((1-\epsilon) c_L(t) I_L(t) + \epsilon c_U(t) I_U(t)\big). \end{aligned} \tag{1}$$

Where $\gamma$ was the recovery rate, $\epsilon$ was the coupling factor between the two groups due to mixing in the same POIs (*33*, *34*), and, $N_L = P_{eff}N$, where $N$ was the population size of the county, and $P_{eff} \in [0,1]$ was the proportion of the population in the lower SES group. The full set of ODEs for each county was,

$$\begin{aligned} \partial_t S_L &= -c_L(t) S_L \lambda_L(t), \\ \partial_t E_L &= c_L(t)(S_L + \sigma W_L)\lambda_L(t) - \alpha E_L(t), \\ \partial_t I_L &= \alpha E_L(t) - \gamma I_L(t), \\ \partial_t R_L &= \gamma I_L(t) - \omega R_L(t), \\ \partial_t W_L &= \omega R_L(t) - \sigma c_L(t) W_L \lambda_L(t), \\ \partial_t S_U &= -c_U(t) S_U \lambda_U(t) \\ \partial_t E_U &= c_U(t)(S_U + \sigma W_U)\lambda_U(t) - \alpha E_U(t), \\ \partial_t I_U &= \alpha E_U(t) - \gamma I_U(t), \\ \partial_t R_U &= \gamma I_U(t) - \omega R_U(t), \\ \partial_t W_U &= \omega R_U(t) - \sigma c_U(t) W_U \lambda_U(t). \end{aligned} \tag{2}$$

Where $\sigma$ is the reduction in susceptibility following a primary infection and $\omega$ is the waning rate of natural immunity.

The *S,E,I,R* disease compartments correspond to numbers of susceptible/naïve, exposed, infectious and recovered/immune (*35*), with subscripts denoting lower or higher SES group. The effective R numbers for an infected in either SES group were,

$$R_{eff,L}(t) = \gamma c_L(t) R_0(t) \left(\frac{\epsilon c_L(t) S_L(t)}{N_L} + \frac{(1-\epsilon) c_U(t) S_U(t)}{N_U}\right),$$
$$R_{eff,U}(t) = \gamma c_U(t) R_0(t) \left(\frac{\epsilon c_U(t) S_U(t)}{N_U} + \frac{(1-\epsilon) c_L(t) S_L(t)}{N_L}\right). \quad (3)$$

Fitting the incubation and recovery rates. The incubation rate and recovery rates were chosen so that

1) The exponential growth rate *r* to basic reproductive number $R_0$ relationship in this model matches that implied by Ferretti et al (*36*). The relationship was calculated using the formula $R_0 = \hat{w}(r)^{-1}$, where $\hat{w}$ was the moment generating function (mgf) of the generation distribution for the transmission model (*37*). Ferretti et al derived a best-fit generation time distribution for SARS-CoV-2 transmission: Weibull(2.8,5.7), and the mgf of the generation time distribution implied by (2) can be solved directly by using Laplace transforms. The importance of matching to *r* to $R_0$ relationship was that *r* is the directly observed quantity from case data, whereas $R_0$ scales directly with reduction in mobility. Therefore, in this study, which is both data-driven and estimates the underlying mobility of the lower SES group, matching the *r* to $R_0$ relationship to an inferred SARS-CoV-2 generation time distribution was important.
2) The mean period between infections and symptom on-set was 5.1 days (*38*), assuming a mean pre-symptomatic period of 2 days (*36*, *39*).

By combining these two criteria we chose incubation rate as $\alpha = 1/3.1$ days$^{-1}$, and the recovery rate $\gamma = 2.4$ days$^{-1}$, see Fig. S2 for agreement between the *r* and $R_0$ relationship for this model and the generation time distribution given by Ferretti et al.

Waning immunity. The *W* disease compartment corresponds to the number of people who have had waning immunity following a natural infection of SARS-CoV-2. Full immunity is assumed to be lost at a rate $\omega$ =1/180 days $^{-1}$, and after loss of full immunity the W-group individuals have a decreased susceptibility to SARS-CoV-2 infection of $\sigma = 0.16$ relative to S-group individuals. There is substantial uncertainty about reinfection, and how transmissible reinfected individuals might be, despite a series of recent studies. We chose the reinfection rate, and subsequent protection from reinfection so that we broadly match these recent studies (Fig. S3). However, we couldn't find any data on whether reinfected individuals are typically as transmissible as people undergoing their first episode of SARS-CoV-2 infection. We defaulted to the maximalist assumption that reinfected individuals are as infectious as the typical person undergoing their primary episode.

Fitting contact rates for visiting POIs for lower and upper SES groups. A key assumption in this paper is that we assumed that there exists a group of people in each Kenyan county who are well described by Google mobility data (*30*). The Google mobility data tracks time spent by users/visitation rate in various settings relative to immediately before the pandemic. We took a fairly simple approach to using this information, we considered the averaged relativity mobility to indoors settings: "retail and recreation", "grocery and pharmacy", "transit stations", and, "workplaces", which we weighted equally. This gave a relative mobility rate to "risky" settings on each day after 20th February 2020, which we assumed was data for fitting $c_U(t)$. We fitted $c_U(t)$ using a simple piecewise linear functional form: $c_U(t) = 1$ for $t \leq$ 15th March 2020 and $t \geq$ 1st November 2020, between these dates we fitted a rapid linear decline in $c_U(t)$ from 15th March to a minimum point on 15th April 2020 $c_U(15th\ April) = 0.56$, and then slower linear growth until 1st November 2020 (Fig S1).

We assumed that the lower SES group followed the same pattern as the Google mobility data: decreasing from 15th March 2020 until 15th April 2020 then increasing back to baseline on 1st November 2020 *except* that the minimum point of the lower SES mobility was treated as an unknown parameter $c_{L,min}$.

Change in transmission rates due to schools being open/closed and introduction of new variants. In addition to the time-varying visitation rates to POIs, we assumed that the basic reproductive number $R_0(t)$ depended on whether (i) schools were open or shut, (ii) whether a, potentially, more transmissible variant than the original wildtype strains had become dominant. Therefore, the reproductive number $R_0(t)$ took three values over the period being simulated (Feb 21st 2020 to Aug 1st 2021):

1. $R_0(t) = R_0$, whilst schools were open (before March 15th 2020 and after Jan 4th 2021) and before Feb 1st 2021. The parameter $R_0$ is the unrestricted reproductive number of the wildtype SARS-CoV-2 strains circulating from the beginning of the pandemic in Kenya.
2. $R_0(t) = \xi_{schools} R_0$, between March 15th 2020 and January 4th 2021 whilst schools were open. $\xi_{schools}$ was the multiplicative effect on $R_0$ of having schools closed.
3. $R_0(t) = \xi_{variant} R_0$, after Feb 1st 2021. $\xi_{variant}$ was the multiplicative effect on $R_0$ of a new variant becoming dominant in February 2021.

The cumulative infection processes. The number of people who would test positive, either as PCR positive, or as seropositive, on each day *n* depended on: 1) the rate of new incidence on each day $s < n$, and, 2) the probability that someone who was infected on day *s* is detectable by either PCR or serology respectively $\tau = n - s$ days later. We calculated the daily incidence rate from the model by including the cumulative infections among the lower and upper SES groups and separating between first infection and all infections (including reinfections) as dummy degrees of freedom in the ODE system (2):

$$\begin{aligned} \partial_t F_L(t) &= c_L(t) S_L \lambda_L(t), \\ \partial_t C_L(t) &= c_L(t)(S_L + \sigma W_L)\lambda_L(t), \\ \partial_t F_U(t) &= c_U(t) S_U \lambda_U(t), \\ \partial_t C_U(t) &= c_U(t)(S_U + \sigma W_U)\lambda_L(t). \end{aligned} \tag{4}$$

Where $F(t)_.$ and $C_.(t)$ were respectively the cumulative first infections, and all infections, by time *t* in each SES group. The daily numbers of new infections on each day *n* among each group and differentiating between first and all infections predicted by the transmission model was,

$$\iota_{n,F,L} = F_L(n+1) - F_L(n) \text{ for each day } n. \tag{5}$$

And similarly for other combinations of SES group and infection episode.

**Observation model**

The underlying transmission of SARS-CoV-2 is not observed, rather we have access to swab tests and serological tests (positive and negative) aggregated by date and county. Therefore, we developed an observation model that connects unobserved daily transmission rates, which depend on the unknown transmission parameters, to the number of individuals in each county who were *detectable* as infected. Then we define parametric distributions for the daily pattern of positive and negative swab and serological tests on each day.

There was substantial day-to-day variation in both reported numbers of positive swab tests and percentage of positive tests among all samples confirmed that day (where negative test data is available). The underlying causes of the high level of day-to-day volatility are probable multiple including variation in daily testing rate, as well as in the settings at which swab test were collected, e.g. at the hospital, from a walk-up testing facility etc, as the focus of Kenyan public health teams has shifted over the course of the epidemic. Because of the substantial day-to-day variation, we use the standard *robust* alternatives to the natural Poisson and Binomial models for count data, the Negative-binomial and Beta-Binomial models respectively (*40*). Moreover, we assumed that the two SES groups had different parameters with respect to the chance of an infection being detected.

Observable infection status in each county. The probability that an infected individual would be determined as having been infected $\tau$ days after infection *if* tested by either a PCR swab test or a serology test was denoted, respectively, $Q_{PCR}(\tau)$ and $Q_{sero}(\tau)$. By combining the underlying infection processes and the delay between infection and observability in our available data sets we find that the number of people who would test positive on each day in each county from either SES group, with a PCR test $(P^+)_{n,L/U}$, and/or a serology test $(S^+)_{n,L/U}$, is,

$$\begin{aligned}(P^+)_{n,L} &= \textstyle\sum_{s=1}^{n-1} \iota_{s,C,L} Q_{PCR}(n-s),\\ (P^+)_{n,U} &= \textstyle\sum_{s=1}^{n-1} \iota_{s,C,U} Q_{PCR}(n-s),\\ (S^+)_{n,L} &= \textstyle\sum_{s=1}^{n-1} \iota_{s,F,L} Q_{sero}(n-s) + p_{FP} S(t),\\ (S^+)_{n,U} &= \textstyle\sum_{s=1}^{n-1} \iota_{s,F,U} Q_{sero}(n-s) + p_{FP} S(t)\end{aligned} \qquad (6)$$

where $t$ was the midpoint of day *n*.

$p_{FP}$ was the false positive rate for the serology assay (see table S1). Underlying equation (6) is an assumption that the PCR test is 100% specific to SARS-CoV-2. Note that we are assuming that *only* the first infection contributes to the serological status of individuals, but that reinfections contribute to PCR status equally to first infections.

Fitting $Q_{PCR}(\tau)$ . We fitted the sensitivity of a PCR swab test on each day $s$ post-symptoms, $Q_\Gamma(s)$, to data on diagnostic accuracy given in Zhou et al (Zhou et al 2020), the fitted functional form for PCR-detectability more than 5 days after infection was:

$$Q_\Gamma(s|\hat{k} = 18.4, \hat{\theta} = 1.1) \text{ for } s \geq 0 \text{ days.} \qquad (7)$$

Where $Q_\Gamma$ was the tail distribution function of a Gamma distribution with fitted shape parameter $\hat{k} = 18.4$ and fitted scale parameter $\hat{\theta} = 1.1$. This aligns with Zhou et al that the median period to become PCR undetectable after symptoms was 20 days with reported interquartile range of 17-24 days (*41*). $Q_{\Gamma}$ doesn't account for the delay between infection and becoming PCR detectable. To account for this delay we assumed that the distribution of delay between infection and maximum detectability followed the same distribution as the delay between infection and onset of symptoms (among those infected individuals who present with COVID-19 symptoms) as reported by Lauer et at (*38*), $timetoonset \sim LogNormal(\widehat{logmean} = 1.64, \widehat{logstd} = 0.36)$. Therefore,

$$Q_{PCR}(\tau) = \sum_{s=0}^{\tau} f_{onset}(s)\, Q_{\Gamma}(\tau - s)\ . \tag{8}$$

Where $f_{onset}(s)$ is the probability of developing symptoms on day $s$ after infection. The true maximum sensitivity of the PCR test in a typical Kenyan setting is absorbed into our observation model via detection probability parameters (see below). $Q_{PCR}$ is displayed in Fig. S3.

<u>Fitting $Q_{sero}(\tau)$.</u> The lag between symptoms and maximum detectability by serological assays has been reported as 21 days in a metastudy of reported diagnostic sensitivities (*42*). We assumed that, given an additional 5 day lag after infection (mean of onset time distribution), the sensitivity of the serological assay increased linearly from 0 at infection to a maximum of 82.5% (*2*) over 26 days.

$$\begin{aligned} Q_{sero}(\tau) &= 0.825\tau/26 \text{ for } 0 < \tau < 26 \text{ days}, \\ Q_{sero}(\tau) &= 0.825 \text{ for } \tau \geq 26 \text{ days}. \end{aligned} \tag{9}$$

$Q_{sero}$ is displayed in Fig. S3. Note that this implementation does not include the possibility of waning levels of detectable antibodies, that is seroreversion (*22*). For comparison in main Fig. 2 we include a forecast of seroprevalence using a simple model of seroreversion,

$$\begin{aligned} Q_{sero}(\tau) &= 0.825\tau/26 \text{ for } 0 < \tau < 26 \text{ days}, \\ Q_{sero}(\tau) &= 0.825(1 - p_w)^{\tau - 26} \text{ for } \tau \geq 26 \text{ days}. \end{aligned} \tag{10}$$

Where $p_w$ is the daily probability of seroreversion if this only occurs on days after the $26^{th}$ day post-infection.

<u>Detection of cases and number of swab tests performed each day.</u> Obviously, our observations depend on the number of swab tests being performed on each day. Because of the fluctuations in testing rate in Kenya, and because of difficulty in asserting the reason for each swab test, we fit to the proportion of positive swab tests on each day whenever positive and negative test results are available (see below). However, we also aim to capture the true underlying detection rate over time; that is the % of all infections who are identified as a case. This detection rate changes with time due to the increasing and decreasing availability of tests, however, the number of tests performed on each day was also somewhat dependent on the demand for tests. Therefore, we expected the number of tests on each day to be correlated with the proportion of tests returning positive, because in periods with high PCR positivity there was also likely to be higher demand for tests to be performed.

We found that the daily number of tests performed across Kenya increased almost monotonically from the beginning of March 2020 until reaching 4000 tests nationwide per day in early July

2020, and afterwards was correlated with the proportion positive over all the tests, as expected (Fig. S4). We interpreted this finding as the testing regime being composed of two main periods: 1) March – June 2020, under-capacity of testing in Kenya when infections were more likely to be unidentified due to simply lacking available swabs, and, 2) July 2020 onwards, Kenyan testing is at maximum capacity and the testing rate responds to demand. We give a simple piece-wise linear form for the relative detection rate (RDR) in Kenya on day $n$,

$$\text{RDR}(n) = \begin{cases} 0, \ n < 14th\ March\ 2020, \\ (n - 14thMarch2020)/120, \ n \geq 14th\ March\ 2020, n < 12th\ July\ 2020, \\ 1, n \geq 12th\ July\ 2020. \end{cases} \quad (11)$$

Negative-binomial model for number of daily positive swab tests. The mean detection rate per PCR-detectable individual per day by swab testing for each SES group ($p_{test,L}$ and $p_{test,U}$), and the clustering factor[1] of the daily detections ($\alpha$), which we point estimate as $\alpha_{PCR} = 0.5$. The observed number of positive swab tests in each county on day n was connected to the underlying detection rate using a negative binomial distribution,

$$\mu_n = RDR(n)(p_{test,L}(P^+)_{n,L} + p_{test,U}(P^+)_{n,U}), \\ (ObsP^+)_n \sim NegBin(\hat{\mu} = \mu_n, \hat{\alpha} = \alpha_{PCR}) \text{ for each day } n. \quad (12)$$

Where $\mu_n$ is the mean number of PCR positives expected by the model accounting for the detection rates by SES group, relative detection rate (which reduces the chance of detecting a case before July 2020)

Beta-binomial model for proportion of daily positive swab tests. We didn't expect the daily swab test results to reflect an unbiased sample of the underlying PCR- detectable fraction of the population. Therefore, we include a relative bias parameter for a PCR-detectable individual being tested relative to a PCR-undetectable individual, for each SES group ($\chi_L$, $\chi_U$), and an effective sample size parameter ($M_{PCR}$; (*40*)). We used a point estimate of $M_{PCR} = 30$. On days where negative swab tests were available, we connect the observable status of the epidemic to the data thus,

$$p_n = (1 - p_U)\frac{\chi_L(P^+)_{n,L}}{(\chi_L - 1)(P^+)_{n,L} + NP_{eff}} + p_U\frac{\chi_U(P^+)_{n,U}}{(\chi_U - 1)(P^+)_{n,U} + N(1 - P_{eff})} \\ (ObsP^+)_{n,c} \sim BetaBin(\widehat{N_s} = N_{PCR,n}, \ \hat{p} = p_n, \widehat{M} = M_{PCR}) \text{ for each day } n. \quad (13)$$

Where $N_{PCR,n}$ is the total number of PCR swab samples collected on day $n$ in the county being fitted and $p_n$ is the proportion of tests performed returning positive expected by the model, accounting for bias in the sampling regime and the possibility that tests occur among the upper SES group with probability $p_U$ independently of the underlying numbers of PCR detectable individuals.

Beta-binomial model for proportion of daily positive serological tests. The reported uncertainty in the maximum sensitivity of serology assay was fairly high: the posterior mean sensitivity was 82.5% (credible interval 69.6-91.2%; (*2*). The posterior uncertainty in the serological sensitivity influenced the confidence the inference method placed on the serological sample data; if the test

[1]Here the clustering coefficient is the inverse of the dispersion parameter $k$, a common alternative parameterisation of the negative binomial distribution.

sensitivity was known to high precision we would treat each day's serological samples as a binomial draw from an underlying proportion of seropositive individuals given by equation (6). Given that the sensitivity of the serological assay was itself an uncertain factor we fitted the posterior uncertainty in the testing sensitivity to a beta distribution: $serological\ sensitivity \sim Beta(\hat{\alpha} = 33.6, \hat{\beta} = 7.13)$. This implied that the appropriate observation model for the number of positive serological samples on day $n$ ($(ObsS^+)_n$), out of the total number of serological samples being collected on day $n$, $N_{sero,n} = (ObsS^+)_n + (ObsS^-)_n$, was a Beta-binomial distribution,

$$(ObsS^+)_n \sim BetaBin(\widehat{N_s} = N_{sero,n}, \hat{p} = \frac{(S^+)_n}{N}, \widehat{M} = M_{sero}). \quad (14)$$

Given an underlying realization of the transmission process the mean number of positive serological samples on day $n$ is $N_{sero,n} \frac{(S^+)_n}{N}$. The "total-count" parameter (*40*), $M_{sero} = \hat{\alpha} + \hat{\beta} = 40.73$, allowed for greater dispersion in the observed seropositive count data than would be allowed by a Binomial model.

#### Parameter inference

As described in **Data description** section we had access to data on daily swab tests (positive and negative) and the KWTRP serological survey daily samples. We grouped the parameters for inference into transmission model parameters ($\theta_{TM}$) and observation model parameters ($\theta_{OM}$) and used Bayesian likelihood-based inference to infer parameters. A challenge with using the linelist data in Kenya for inference of transmission was that the metadata concerning the reason for receiving a swab test, the levels of symptoms of people who tested positive, and their healthcare outcomes were often missing. Overall, more than 90% of the people who tested positive in Kenya, and for whom we have a description of their symptoms, reported no symptoms (asymptomatic). Therefore, unlike model-based inference for COVID-19 transmission in high-income countries we didn't use severe outcomes such as hospitalisation or death as data sources for inference, e.g. (*9*), because this data was unreliable. Instead, we concentrated on fitting to the proportion positive of daily swabs test and serological tests jointly with detection rate of cases.

We use the Bayesian inference to infer a joint posterior distribution for the unknown parameters (both transmission-based parameters and observation-based parameters) for each county. We describe the three main ingredients for our Bayesian approach below: 1) the log-likelihood function for the data given a set of parameters, 2) the county-specific hierarchy of prior distributions for the parameters, and, 3) the Markov-chain Monte Carlo method used to draw parameter sets from the posterior distribution.

Log-likelihood function. The observation model gives the following log-likelihood function for the unknown parameters $\theta = (\theta_{TM}, \theta_{OM})$ given the sampling data for a county, $\boldsymbol{D} = \{(ObsS^+)_n, (ObsS^-)_n, (ObsP^+)_n, (ObsP^-)_n\}_{n=1,2,3,\ldots}$ :

$$l(\theta) = \sum_n ln\, f_{NB}((ObsP^+)_n | \hat{\mu} = \mu_n, \hat{\alpha} = \alpha_{PCR}) + \sum_{n \in negs} ln\, f_{BB}((ObsP^+)_n | \widehat{N_s} = N_{PCR,n}, \hat{p} = p_n, \widehat{M} = M_{PCR}) + \sum_n ln\, f_{BB}(\,(ObsS^+)_n | \widehat{N_s} = N_{sero,n}, \hat{p} = \frac{(S^+)_{n,c}}{N}, \widehat{M} = \widehat{M}_{sero}). \quad (15)$$

Where $f_{NB}$ and $f_{BB}$ are, respectively, the probability mass functions for the negative binomial and beta-binomial distributions, as described in the observation model subsection. The index set of days in $negs$ covered days where PCR negative tests were available in the county being fitted. The first day where samples were included in the log-likelihood calculation was 12th April 2020, due to testing being even more irregular before that date, and the last day was 27th April 2021.

County-specific hierarchy of priors. As described in the main text, when designing priors for Bayesian inference we, first, grouped the counties by socio-economic and epidemiological factors using an unsupervised machine learning technique. The socio-economic/epidemiological factors are described and were generated in Macharia et al (*27*), and included factors such as county rates of obesity, informal settlement habitation, population density and access to major cities. We used ordered leaf clustering by centred and standardized $L_1$ (Manhattan) distance over all factors to group the 47 counties and identified four groups of similar counties (Fig S5). Nairobi and Mombasa were sufficiently distinct from other counties that they formed their own singlet groupings. There were two further county groupings evident from the unsupervised learning, which corresponded to predominantly remote and rural counties and to counties with partial urbanization and greater connectivity to the main Kenyan cities. The full county list of the identified groups was:

1) The capital city **Nairobi** (also a county) and,
2) The second city **Mombasa** (also a county)
3) Semi-urban/semi-rural counties that either contain significant sized cities and/or neighbour Nairobi county: **Baringo, Bomet, Bungoma, Busia, Elgeyo Marakwet, Embu, Homa Bay, Kakamega, Kajiado, Kericho, Kiambu, Kirinyaga, Kisii, Kisumu, Kitui, Laikipia, Machakos, Makueni, Meru, Migori, Murang'a, Nakuru, Nandi, Nyamira, Nyandarua, Nyeri, Siaya, Taita Taveta, Tharaka Nithi, Trans Nzoia, Uasin Gishu, Vihiga**.
4) More rural counties and/or more remote counties: **Garissa, Isiolo, Kilifi, Kwale, Lamu, Mandera, Marsabit, Narok, Nyeri, Samburu, Tana River, Turkana, Wajir, West Pokot**

We also distinguished the counties by the number of serological tests in the KWTRP serological surveillance trial rounds 1 and 2 that had been performed on people living in them. Serology test data was critical for identifying parameters involved in the detection of infections in each county by giving an estimate of the true population exposure at different time points. 11 counties had notably more serological tests than the 36 other counties: **Embu, Kilifi, Kisii, Kisumu, Kwale, Mombasa, Nakuru, Nairobi, Nyeri, Siaya, Uasin Gishu** (Fig S5).

For the 11 counties with higher numbers of serological tests we assigned priors that encoded our *a priori* beliefs about the epidemic in Kenya. The prior distributions for $\epsilon$, $c_{L,min}$, $\chi_L$, $\chi_U$ were the same for each county reflecting our a priori beliefs. We had 1) strong confidence that mixing between SES groups would be assortative, 2) weak confidence that the lower SES group would be able to reduce their mobility less than the upper SES group, 3) moderate confidence that the daily detected proportion PCR tests being positive would be higher than the actual proportion of the population PCR detectable, with this bias being higher among the lower SES group who were believed would be less likely to seek a test when asymptomatic compared to individuals in the upper SES group, and, 4) moderate confidence that new variants dominating transmission from February 2021 onwards were about 50% more transmissible:

- $\epsilon \sim Beta(\hat{\alpha} = 45, \hat{\beta} = 5)$.
- $c_{L,min} \sim Beta(\hat{\alpha} = 8, \hat{\beta} = 2)$.
- $\chi_L \sim \Gamma(\hat{k} = 10, \hat{\theta} = 4.5/10)$.
- $\chi_U \sim \Gamma\left(\hat{k} = 10, \hat{\theta} = \frac{1.5}{10}\right)$.
- $\xi_{variant} \sim \Gamma\left(\hat{k} = 10, \hat{\theta} = \frac{1.5}{10}\right)$.

The county group specific priors for the 11 counties with higher numbers of serological tests were:

- **Nairobi and Mombasa.**
  - $R_0 \sim \Gamma(\hat{k} = 3, \hat{\theta} = \frac{2.5}{3})$.
  - $E(0) \sim \Gamma(\hat{k} = 3, \hat{\theta} = 100/3)$.
  - $p_{test,L} \sim \Gamma\left(\hat{k} = 3, \hat{\theta} = \frac{10^{-4}}{3}\right)$.
  - $p_{test,U} \sim \Gamma\left(\hat{k} = 3, \hat{\theta} = 5 \times \frac{10^{-4}}{3}\right)$.
  - $p_U \sim Beta(\hat{\alpha} = 40, \hat{\beta} = 60)$.
  - $P_{eff} \sim Beta(\hat{\alpha} = 35, \hat{\beta} = 15)$.
- **Semi-urban/semi-rural counties.**
  - $R_0 \sim \Gamma(\hat{k} = 3, \hat{\theta} = \frac{2}{3})$.
  - $E(0) \sim \Gamma(\hat{k} = 3, \hat{\theta} = 1/3)$.
  - $p_{test,L} \sim \Gamma\left(\hat{k} = 3, \hat{\theta} = 2 \times \frac{10^{-5}}{3}\right)$.
  - $p_{test,U} \sim \Gamma\left(\hat{k} = 3, \hat{\theta} = \frac{10^{-4}}{3}\right)$.
  - $p_U \sim Beta(\hat{\alpha} = 80, \hat{\beta} = 20)$.
  - $P_{eff} \sim Beta(\hat{\alpha} = 95, \hat{\beta} = 5)$.
- **Rural counties.**
  - $R_0 \sim \Gamma(\hat{k} = 3, \hat{\theta} = \frac{1.5}{3})$.
  - $E(0) \sim \Gamma(\hat{k} = 3, \hat{\theta} = 0.1/3)$.
  - $p_{test,L} \sim \Gamma\left(\hat{k} = 3, \hat{\theta} = 1 \times \frac{10^{-5}}{3}\right)$.
  - $p_{test,U} \sim \Gamma\left(\hat{k} = 3, \hat{\theta} = 5 \times \frac{10^{-5}}{3}\right)$.

- $p_U \sim Beta(\hat{\alpha} = 90, \hat{\beta} = 10)$.
- $P_{eff} \sim Beta(\hat{\alpha} = 95, \hat{\beta} = 5)$.

The county group specific priors were based on the view that although the most *a priori* likely possibility was that most counties had very few infected individuals on 21st February, and that we might expect the unknown numbers of exposed people to be concentrated in cities. We had moderate a priori confidence that the overall detection rate among the upper SES would be around 5% of infections, and about 1% among the lower SES group. We had strong confidence that the inferred community size of the upper SES group would be higher in cities rather than other counties.

For the other 36 counties with less serological testing, we used the same county-group specific priors except that the PCR positivity bias parameters ($\chi_L, \chi_U$) and daily detection rates of positives ($p_{test,L}, p_{test,U}$) had priors derived from the inferred posteriors of counties among the 11 with higher numbers of serology tests. This was an approximation to a hierarchical Bayesian formulation of a joint likelihood for all 47 Kenyan counties; we fitted gamma distributions to the pooled posterior draws for these parameters over the semi-urban/semi-rural counties with larger numbers of serological tests (**Embu, Kisii, Kisumu, Nakuru, Nyeri, Siaya, Uasin Gishu**) and the rural counties with larger numbers of serological tests (**Kilifi** and **Kwale**). These fitted priors were used for parameter inference.

MCMC draws. We used Hamiltonian MCMC with NUTS *(20) to* perform Bayesian inference by drawing 2,000 samples from the posterior distribution,

$$\theta^{(k)} \sim P(\theta|D) \propto exp\,(l(\theta))\,P(\theta), \text{ for } k = 1,2,3,\ldots \quad (16)$$

for each county using the NUTS-HMC sampler implemented by the Julia language package *dynamicHMC.jl*. Solving the likelihood function for a proposed value of $\theta$ involved solving the ODE system (2), we used the highly performant DifferentialEquations.jl package for ODE solutions (*43*). The HMC method required a log-likelihood gradient, $\nabla_\theta l$, which, for our use-case of a small ODE system with a low number of parameters, was most efficiently supplied by forward-mode automatic differentiation implemented by the package *ForwardDiff.jl*.

The MCMC chain converged for each county (all MCMC chains and MCMC diagnostics can be accessed through the linked open code repository). The posterior mean (and 95% CIs) for each parameter can be found in supplementary data: Data S?

Back-calculation and forecasting for Kenyan counties

The parameter draws from the posterior distribution defined the uncertainty in our model nowcasts and forecasts for each county, since the underlying transmission model was deterministic. Therefore, posterior distributions for epidemic quantities were created by simulating the epidemic for each $\theta^{(k)}, k = 1,2,\ldots$ posterior draws. We used this technique for both back-calculating estimates of quantities that were unobserved during the data collection period, such as the daily infection rate in each SES group and the seropositivity rates after round 2 of the serological survey finished, and, forecasting ahead of the data collection period by assuming that the parameters remained unchanged in May-July 2021.

When forecasting the number of positive tests that would occur on days in the future, we combined a forecast of the proportion positive expected on each day, for each posterior draw of a parameter set, with a forecast of the number of tests that would be performed on each day. We noted that the number of tests performed was not independent of the proportion positive, and for each county fitted a simple linear model

$$N_{PCR,n} = \beta_0 + \beta_{prop} \frac{(ObsP^+)_n}{N_{PCR,n}} + \epsilon_n \,. \tag{17}$$

For days $n$ that cover the most recent 60 days of available test data ($\epsilon_n$ are iid normal errors).

Inference of mortality rate per infection

The commonly used infection fatality ratio (IFR) by age estimates from Verity et al (*44*), weighted by the Kenyan population distribution given by the 2019 census, implied a basic IFR prediction in Kenya of $IFR_{verity} = 0.264\%$; that is 264 deaths per 10,000 infections. This assumed a uniform attack rate across age-groups in Kenya.

We used the posterior predictions of the underlying daily infections in Kenya counties to infer a crude infection fatality ratio ($IFR_{crude}$) for each Kenyan county. The lag between infection and death, for those infected individuals who die, was defined as the convolution of three time duration distributions:

1. The duration of time between infection and symptoms (days), which we assumed was distributed $LogNormal(\widehat{log\mu} = 1.64, \widehat{logstd} = 0.36)$ (*38*).
2. The duration of time between initial symptoms and severe symptoms (days), sufficient to seek hospitalisation, which we assumed was distributed $U(1,5)$ (*45*).
3. The duration of time between severe symptoms and death estimated from UK hospital data (*9*). This was an empirical distribution with mass function $p_{HD}$.

We discretized the first two distributions to give probability mass functions $p_{IS}$ for the number of days between infection and symptoms, and $p_{SH}$for the number of days between symptom onset and severe symptom onset. The probability mass function for the (discrete) number of days between infection and death, for those who died, $p_{ID}$, was given as a discrete convolution over these probability mass functions:

$$p_{ID}(\tau) = p_{IS} * p_{SH} * p_{HD}(\tau) \text{ for the probability that death occurs } \tau \text{ days after infection.} \tag{18}$$

We didn’t use mortality data for inference of parameters; therefore, we estimate mortality detection rates from back-calculation of the underlying daily incidence rate. Because of the possibility of substantial under-reporting of mortality due to COVID-19 in Kenya, which may differ across socio-economic groups, we jointly fit a mortality-detection rate to both SES groups in each county $\mu = (\mu_L, \mu_U)$. We used maximum likelihood estimation to infer mortality-detection rates:

$$\hat{\mu} = argmax_{\mu} \sum_t ln\, f_{poi}(D(t) | [(\mu_L \overline{\iota_L} + \mu_U \overline{\iota_U}) * p_{ID}](t)). \tag{19}$$

Where $D(t)$ were the number of deaths attributed to COVID-19 on each day $t$, $\overline{\iota_L}$ and $\overline{\iota_U}$ were the posterior mean back-calculations for the daily rate of new infections among the lower and upper SES groups, which were convolved with a probability kernel $p_{ID}$ for the lag in days between infection and death.

**Further evidence to support the two social group model**

Comparison to modelling the first wave. A previous model for the first wave in Kenya used a similar methodology, but rather than using two SES groups each county was modelled as forming a single transmission group with an effective population size to account for heterogeneity in transmission and with R(t) fitted daily rather than assuming the simple parametric form used in this paper (*6*). The one-group model was successful in capturing the dynamics of the first wave in Kenya, and is close agreement with the model presented in this study over the course of the first wave in Kenya, for example, the one-group model predicted 43.3% (CI 35.3%-49.5%) by 30th September (*6*) whereas the posterior mean estimate for overall exposure in Nairobi for the two group model was 40.0%. However, the one-group model failed to explain the second wave in Kenya without large increasing in R(t) relative to the February 2020 estimates, with these increases starting months before VOCs like B.1.1.7 were first detected in Kenya (Fig S6). These large increases in R(t) for the one-group model derived from the model having a single detection rate for all infecteds, whereas the two-group model favours an explanation where infections in the lower SES group are detected at lower rates than infections in the upper SES group.

Randomised seroprevalence survey in Nairobi. The model that we present in our current paper proposes separate SES groups with infections in the upper SES group being both delayed and more densely sampled compared to the lower SES group. An obvious alternative explanation is that the serology data used to fit the model was an over-estimate of exposure, possibly due to the underlying blood donor serology data being heavily biased in favour of the detection of seropositive individuals. However, the overall model prediction of seropositivity in Nairobi was in reasonable agreement with the overall seroprevalence random selection of the Nairobi population performed in the first half of November 2020 (*46*). The agreement between the model prediction and the randomized serological surveys was even better for the central age groups (ages 10-60), whereas the overall seropositivity (including <10 year olds and > 60 year olds) was lower. This may reflect model bias in that we calibrate to blood donor serology, which limits to donors in the 15-65 age interval. However, first, the predictive error was not substantial (< 10% seropositivity prediction error), and, second, it is not clear if this bias will persist after general reopening of schools in Kenya (Fig S7).

The randomised seroprevalence survey also reported a breakdown by subcounty *(46).* For comparison to model predictions we compared some exemplar sub-counties to predictions for lower and SES group model predictions of seroprevalence. The Kibera sub-county, containing the Kibera slum area, ranked highest out of 17 Nairobi sub-counties for both poverty and density of informal settlement and we used the seroprevalence of this county as an exemplar of the lower SES group. No Nairobi sub-county was lowest ranked for both poverty and informal settlement density so as the upper SES group exemplar we used the seroprevalence over three sub-counties: Dagoretti South (12th for poverty, 15th-17th for informal settlement), Embakasi East (15th-17th for poverty, 12th for informal settlement) and Roysambu (14th for poverty, 13th for informal settlement). Poverty and informal settlement density data was sourced from Macharia et al *(27).*

The agreement between the exemplar sub-county unadjusted seroprevalences and the model predictions were also reasonable, which suggested that segregating the population by SES status was a reasonable approach to capturing the wide disparity in seroprevalence between Nairobi sub-counties reported in *(46)*.

Data on public and private hospital admissions. We had imperfect data on hospital admissions in Nairobi, derived from the Kenyan national linelist, and did not use admissions to fit the model. However, in the Kenyan capital Nairobi we had a subset of cases with PCR swab test confirmation *and* with metadata on admission to a named health facility. During the first wave we note that most of the identified admissions occurred in public hospitals, which would be selected by people in the lower SES group, whereas in the second wave private health facilities were more frequently reporting admissions (Fig S8).

#### Supplementary Text

Notation for distributions used in this study

In this study, we have used a number of parameter symbols that are also the most commonly used symbols for various common parametric distributions. Moreover, these parametric distributions are used in the underlying analysis frequently with their distribution parameters defined as functions of underlying transmission model states. To reduce misunderstanding reserve symbols with "hats" as referring to the parameters of a parametric distribution and use "=" to refer to the value of the parameter. Find below the choice of parametrization for the parametric distributions used in the study:

- Exponential distribution. $Exp(\hat{\mu} = \mu)$, with mean $\hat{\mu}$.
- Gamma distribution. $\Gamma(\hat{k} = k, \hat{\theta} = \theta)$, with shape parameter $\hat{k}$ and scale parameter $\hat{\theta}$.
- LogNormal distribution. $LogNormal(\widehat{logmean} = logmean, \widehat{logstd} = logstd)$, with log-mean parameter $logmean$ and log-standard deviation $logstd$.
- Negative binomial distribution. $NegBin(\hat{\mu} = \mu, \hat{\alpha} = \alpha)$, with mean $\hat{\mu}$ and clustering factor (inverse dispersion parameter) $\hat{\alpha}$.
- Beta distribution. $Beta(\hat{\alpha} = \alpha, \hat{\beta} = \beta)$, with shape parameters $\hat{\alpha}, \hat{\beta}$ (mean $\hat{\alpha}/(\hat{\alpha} + \hat{\beta})$).
- Beta-binomial distribution. $BetaBin(\widehat{N_s} = N_s, \hat{p} = \mu, \widehat{M} = M)$, with number of samples $\widehat{N_s}$, marginal probability per draw $\hat{p}$, and effective sample size $\widehat{M}$ (see for details of this slightly unusual parameterization).
- Poisson distribution. $Poisson(\hat{\mu} = \mu)$, with mean $\hat{\mu}$.

**Fig. S1.**

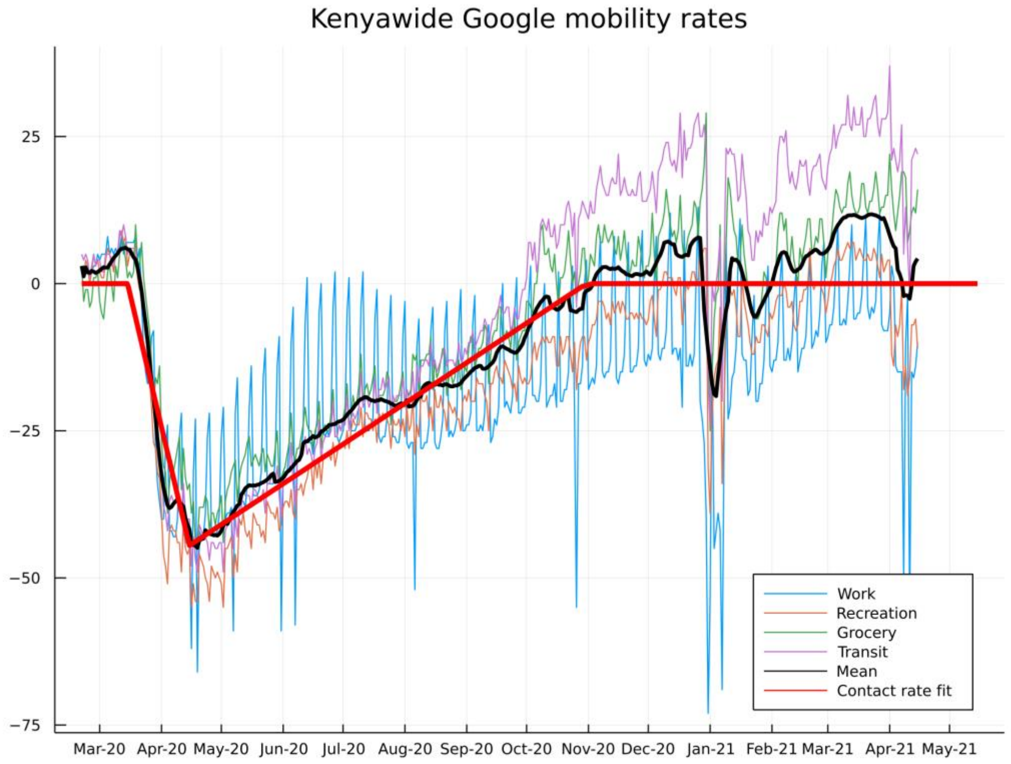


Fig S1: **Google mobility trends for Kenya**. The curves show mobility rates of smartphone users to Google defined setting categories, "retail and recreation", "grocery and pharmacy", "transit stations", and, "workplaces". Also, shown is the 7-day moving average of the mean over these settings (*black curve)* and the approximation for the contact rate used for the upper SES group in this paper (*red curve)***.**

**Fig. S2.**

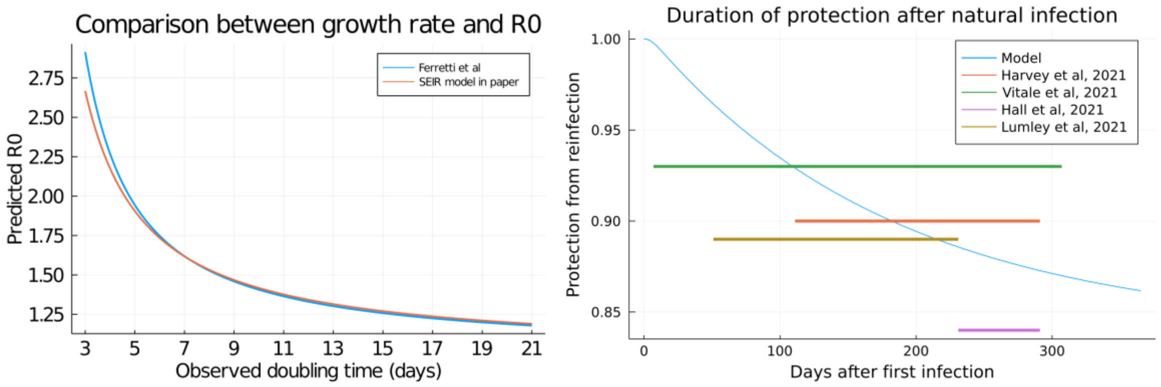


Fig S2: **Comparisons between model structure and SARS-CoV-2 observation: relationship between exponential growth rate and reproductive number, and, duration of protection against natural infection.** *Left:* Comparison of relationship between doubling time and $R_0$ for the model used in this paper and a data-derived generation distribution for SARS-CoV-2 *(36)*. The relationship between the exponential growth rate *r* and the reproductive number was, $R_0 = \frac{1}{\widehat{W}(r)}$, where $\widehat{W}(r)$ was the moment generating function of the generation time distribution evaluated at *r*. *Right*: The model prediction for relative protection from reinfection due to a natural compared to a fully naïve individual (reduction in probability of infection per challenge) as a function of time since infection. Colored bars are levels of protection reported by four studies *(13-16)*, horizontal width indicates the period after infection that the study reports this level of protection.

**Fig. S3.**

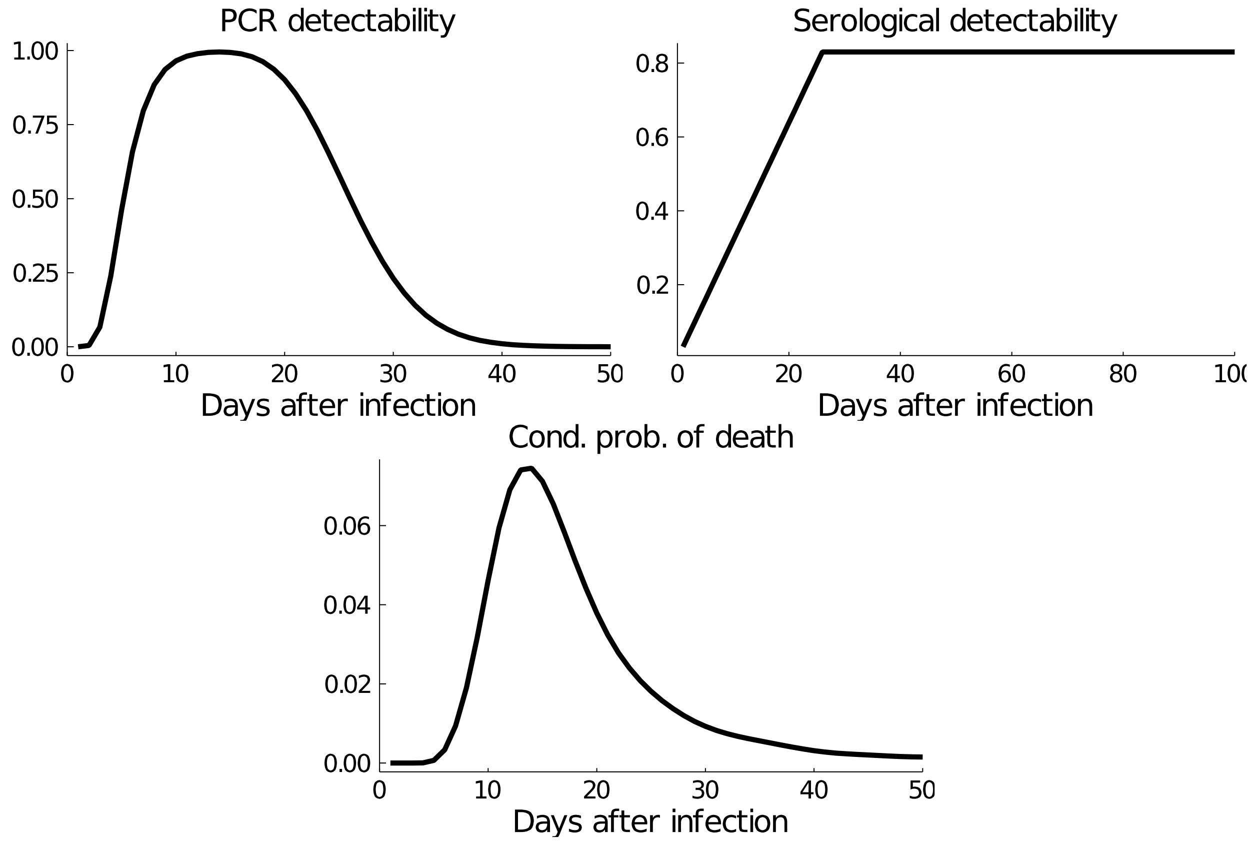


Fig S3: **Distributions of time between infection and observable events for those infected.**

The time-since-infection dependent probability of being detectable by a PCR test *(top left*) or serology test *(top right)* used in this paper. The distribution of time between infection and death, conditional on a death outcome *(bottom)* used in this paper.

**Fig. S4.**

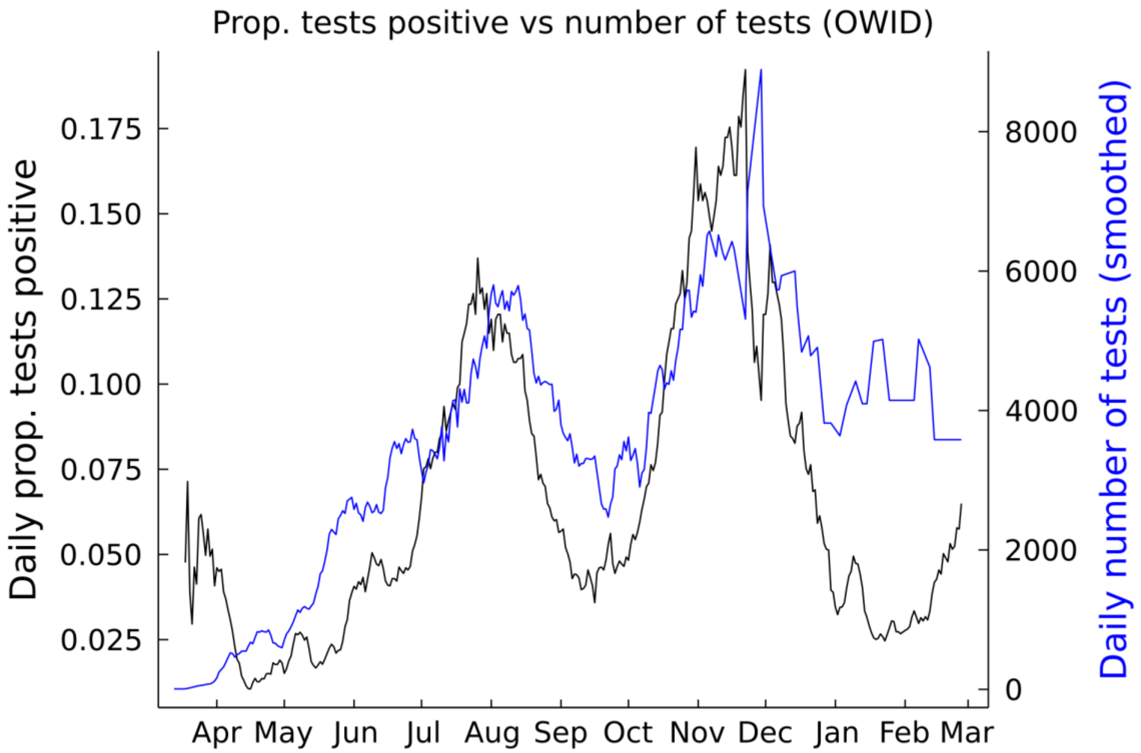


Fig S4: **Trend in daily testing in Kenya March 2020-March 2021.** The number of negative tests was not available for every county in this period; therefore, we default to Kenya Ministry of Health tweets of national testing rates (collated by ourworldindata.org). *Left axis*: The rolling 7-day average positivity rate per test. *Right axis*: The rolling 7-day average daily number of tests performed.

**Fig. S5.**

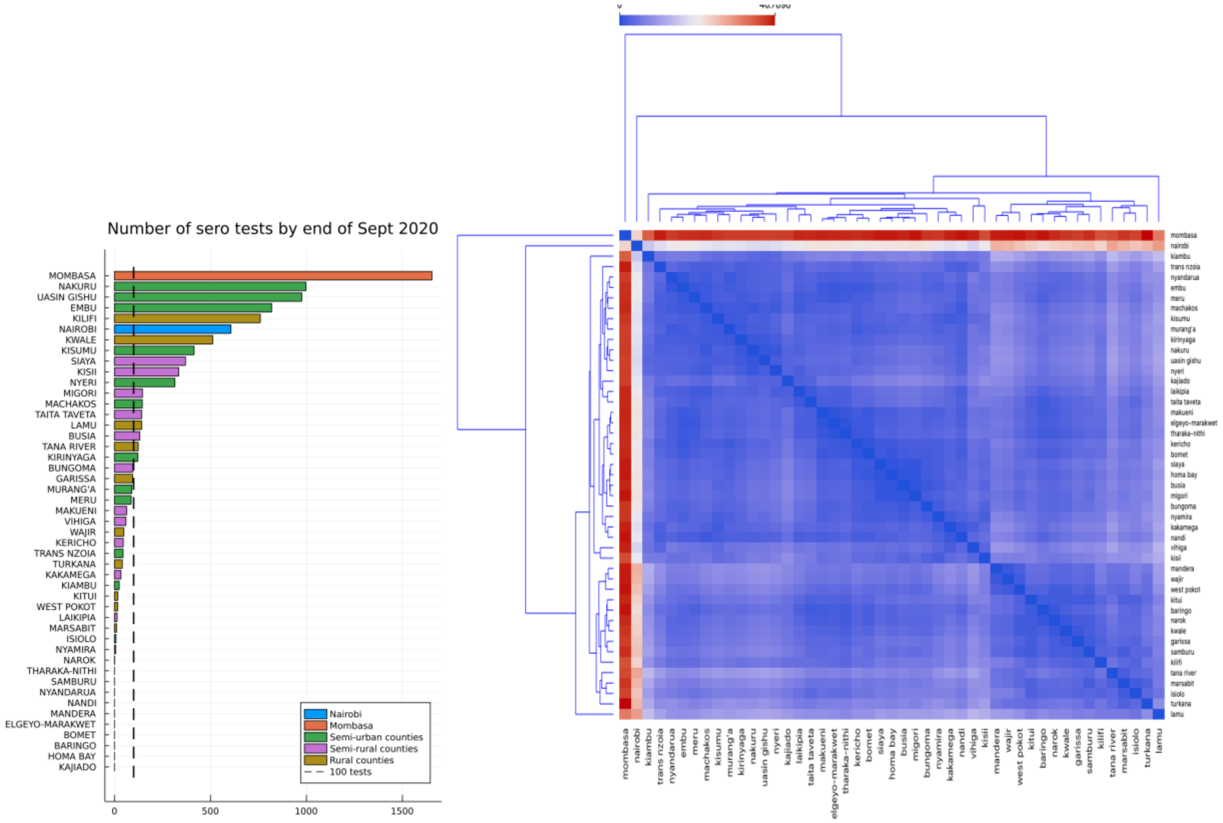


Fig S5: **Grouping variables for Kenya counties.** *Left:* The number of serology tests available by county up until September 2020. Counties with as many serology tests as Nyeri county or more used moderately informative priors for detection rate parameters, counties with less serology tests used fitted priors for detection rate parameters. *Right*: Distance matrix (normalized $L_1$ distance) used to group counties into four categories: Nairobi, Mombasa, semi-urban/semi-rural and rural.

**Fig. S6.**

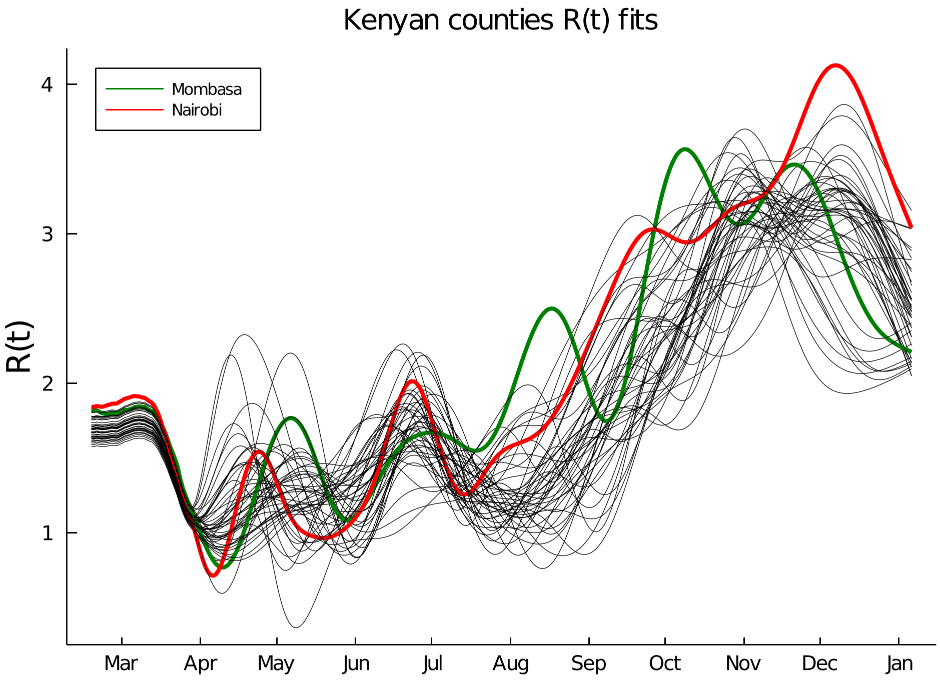


Fig S6: **Fitted R(t) from a one-group county-based transmission model.** The estimated R(t) values for all 47 counties fitted using a one-group per county model for SARS-CoV-2 transmission in Kenya *(6)*. The one-group model can only fit the available data in 2020 by postulating big increases in R(t) above baseline estimates from September 2020 (before the detection of any VOCs in Kenya). The model presented in this paper favoured an explanation where the lower SES group transitioned to high population exposure, then the upper SES group combined with under-sampling among the lower SES group.

**Fig. S7.**

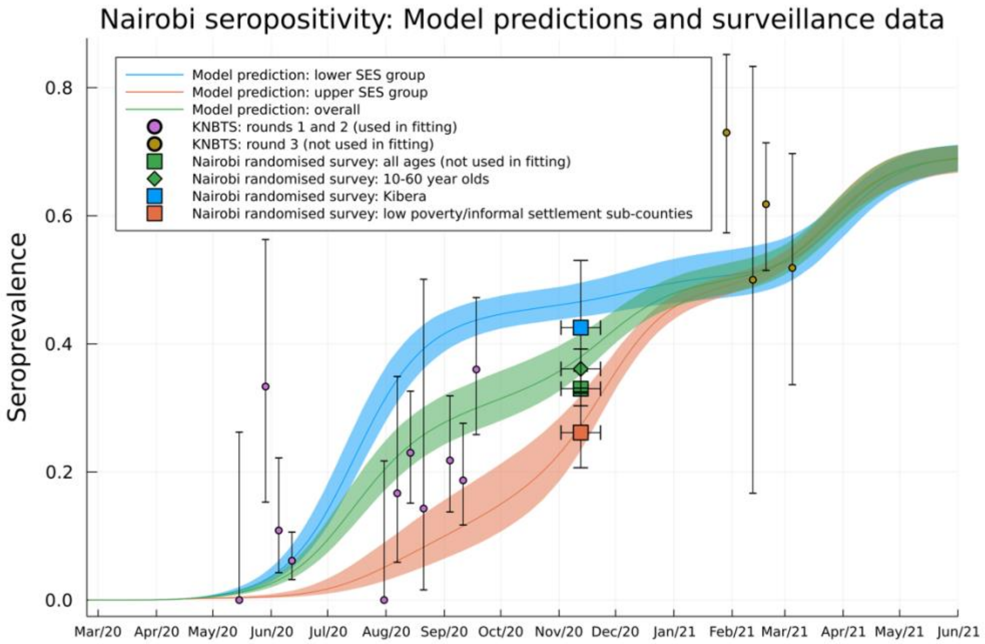


Fig S7: **Additional seroprevalence surveys for SES group dynamics in Nairobi.** The SES specific, and overall, seropositivity prediction from the model (*curves*) with round one and two KNBTS blood donor weekly seroprevalence (*purple dots*) which was used to fit the model for Nairobi. Also shown are data the following seroprevalence surveys, not used to fit the model and displayed for comparison and model validation: 1) Round three KNBTS blood donor weekly seroprevalence (*brown dots*), and 2) a randomized survey of all Nairobi households *(46)*. The randomized household surveys are presented as over all age groups (*squares*) and over 10–60-year-olds (*diamonds*). Also shown the seroprevalence for the Kibera sub-county, the Nairobi sub-county with the highest rates for poverty *and* informal settlement density (*blue square; 27*), and, the combined seroprevalence for Dagoretti South, Embakasi East and Roysambu sub-counties, which are among the Nairobi sub-counties with lowest rates for poverty *and* informal settlement density (*red square; 27*). Bars represent either 95% Jeffery's confidence intervals (vertical bars) or the duration of the survey (horizontal bars). Background shading indicates 95% confidence intervals from the posterior distribution of parameters.

**Fig. S8.**

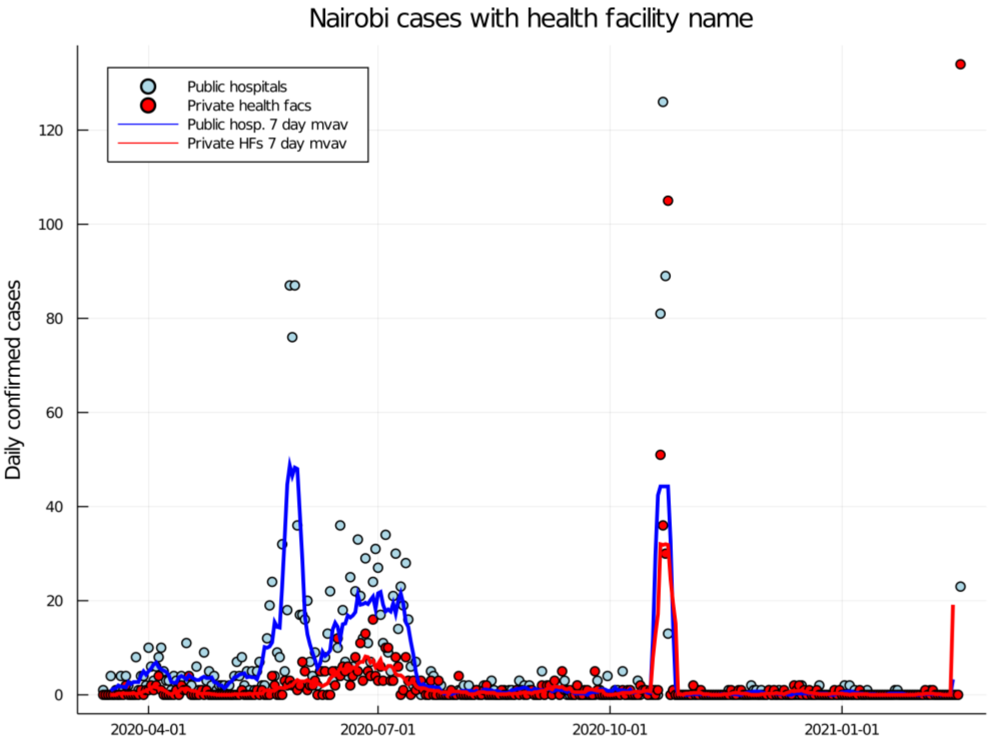


Fig S8: **Confirmed cases in Nairobi by public vs private health facility during first two waves.** Daily confirmed positive swab tests in Nairobi with metadata about admission to a health facility grouped by public hospital (*blue dots; blue curve* is 7-day moving average) and private hospitals/health facilities (*red dots; red curve* is 7-day moving average).

#### Table S1.

| Parameters and variables | Value |
|---|---|
| **County specific transmission model parameters** | |
| Number of susceptible people at time t in lower and upper SES groups, $S_{L/U}(t)$ | Dynamic |
| Number of latently infected people at time $t$ in lower and upper SES groups, $E_{L/U}(t)$ | Dynamic |
| Number of infectious people at time $t$in lower and upper SES groups, $I_{L/U}(t)$ | Dynamic |
| Number of recovered and immune people at time $t$ in lower and upper SES groups, $R_{L/U}(t)$ | Dynamic |
| Number of susceptible people at time t in lower and upper SES groups, who have had a previous infection, $W_{L/U}(t)$ | Dynamic |
| Initial numbers in each infection state on 21st Feb $S_{L/U}(0), E_{L/U}(0), I_{L/U}(0), R_{L/U}(0), W_{L/U}(0)$. | $\boldsymbol{E_L(0) = E_U(0)}$ **was inferred from data.** $I_{L/U}(0) = R_{\frac{L}{U}}(0) = W_{\frac{L}{U}} = 0.$ $S_L(0) = NP_{eff} - E_L(0)$ $S_U(0) = N(1 - P_{eff}) - E_U(0)$ Where the county population size N is as reported in the 2019 Kenyan census. |
| Mean latent (infected but not infectious) period $1/\alpha$ | 3.1 days. The mean incubation period (*38*) was reduced by two days of pre-symptomatic transmission *(39)* to give a latency period. |
| Mean infectious period $1/\gamma$ | 2.4 days. Chosen to fit the exponential growth rate *r* to reproductive number R relationship of a fitted SARS-CoV-2 generation duration *(36)* |
| Baseline reproductive number, $R_0$. | **Inferred from data** |
| Population proportion in lower SES group, $P_{eff}$. | **Inferred from data** |
| Mobility of upper SES group, $c_U(t)$. | Piecewise linear fit to Google mobility data. $c_U(t) = 1$ for $t \leq 15^{th}$ March 2020 and $t \geq 1^{st}$ November 2020. Linear trend decreasing to, and then increasing from, $c_U(15th\ April) = 0.56$. |
| Mobility of lower SES group, $c_L(t)$. | Piecewise linear. $c_L(t) = 1$ for $t \leq 15^{th}$ March 2020 and $t \geq 1^{st}$ November 2020. Linear trend decreasing to, and then increasing from, $\boldsymbol{c_L(15th\ April)}$ **which was inferred from data.** |
| Proportion of secondary infections within same SES group, $\epsilon$. | **Inferred from data** |
| Mean period of complete protection after recovery, $1/\omega$. | 180 days, point estimate based on reinfection studies *(15-18)*. |
| Relative susceptibility compared to naïve susceptibles after loss of complete protection after first infection episode, $\sigma$. | $\sigma = 0.16$, point estimate based on reinfection studies *(15-18)*. |
| Relative effect on transmission of having schools closed, $\xi_{schools}$. | **Inferred from data** |

| | |
|---|---|
| Relative effect on transmission post 1st February 2021 due to introduction of variants of concern, $\xi_{variant}$. | **Inferred from data** |
| **County-specific observation model parameters and data** | |
| Number of people who would test PCR positive on day *n* in lower/upper SES group, $(P^+_{L/U})_n$. | Dynamic |
| Number of people who tested PCR positive on day *n*, $(ObsP^+)_n$. | Data |
| Number of people who would test as seropositive on day *n* in lower/upper SES group, $(S^+_{L/U})_n$. | Dynamic |
| Number of people who tested as seropositive on day *n*, $(ObsS^+)_n$ . | Data |
| Probability that an infected individual would test PCR positive on day $\tau$ after infection, $Q_{PCR}(\tau)$ | $Q_{PCR}(\tau) = f_{onset} * Q_{\Gamma}(\tau)$, where $Q_{\Gamma}$ was the tail function of a gamma distribution fitted to data given in (*41*), and $f_{onset}$ is the probability function of onset of symptoms post-infection (*38*). |
| Probability that an infected individual would be detectably seropositive on day $\tau$ after infection, $Q_{sero}(\tau)$ | $Q_{sero}(\tau)$ is linearly increasing over 26 days to saturate at 82.5% sensitivity, based on report delay in seroconversion (*42*) and maximum sensitivity of serological assay (*2*). |
| Daily rate of PCR-positive individual in lower SES group receiving a swab test, $p_{test,L}$. | **Inferred from data** |
| Daily rate of PCR-positive individual in upper SES group receiving a swab test, $p_{test,U}$. | **Inferred from data** |
| Clustering coefficient of daily PCR tests, $\alpha_{PCR}$. | Point estimate: $\alpha_{PCR} = 0.5$. |
| Relative bias in favour of selecting a PCR positive individual for swab testing among lower SES group (used when negative tests are available) ($\chi_L$) | **Inferred from data** |
| Relative bias in favour of selecting a PCR positive individual for swab testing among upper SES group (used when negative tests are available) ($\chi_U$) | **Inferred from data** |
| Proportion of daily swab tests among upper SES group (used when negative tests are available) ($p_U$) | **Inferred from data** |
| Effective sample size/clustering coefficient of daily PCR tests (used when negative tests are available), ($M_{PCR}$). | Point estimate: $M_{PCR} = 30$. |
| **County-specific detected fatality rate parameters** | |
| Chance of fatality and being determined as due to COVID-19 disease per infection in the lower and upper SES groups, $\mu_{L/U}$. | **Inferred from data** |
| Probability mass function of delay lag between infection and death for those who die, $p_{ID}(\tau)$. | Derived as a convolution over the lag from infection to symptom onset (*38*), the lag from symptoms to hospitalisation/severe symptoms (*45*), |

| | |
|---|---|
| | and the lag between severe symptoms and death (*9*). |

Table S1: **Dynamic and observational model variables and parameters.** “Dynamic”, means that the variable was an output of the transmission and observation model for the county.

**Data S1. (Separate file)**

**Inferred parameters, SES group-specific fatality-detection estimates, and population exposure by 1st June.** The posterior mean and 95% credible intervals for each model parameter (both transmission and observation models), maximum likelihood estimate for infection fatality-detection ratio, and posterior mean and 95% credible intervals for fraction of population exposed in each county.

**Data S2. (Separate file)**

**The number of positive, and negative where available, PCR-confirmed swab tests for each county by date of sample collection (21st Feb to 27th May).** A "-17" in the negative swab entry indicates that negative swab tests were not available for that county on that date.

**Data S3. (Separate file)**

**The number of positive and negative serological results for each county by date of sample collection (21st Feb to 31st March).**

**Data S4. (Separate file)**

**The number of deaths with a PCR-confirmed swab test for each county by recorded date of death (21st Feb to 27th May).**
